# Advancing coupled behavioural-epidemic models: An interdisciplinary framework for the collection of empirical data

**DOI:** 10.1101/2025.10.07.25337410

**Authors:** Vittoria Offeddu, Elisabetta Colosi, Lorenzo Lucchini, Laura Pasqua Leone, Chiara Chiavenna, Duilio Balsamo, Elena D’Agnese, Francesco Bonacina, Maria Cucciniello, Filippo Trentini, Alberto Aleta, Piero Manfredi, Yamir Moreno, Márton Karsai, Vittoria Colizza, Júlia Koltai, Alessia Melegaro

## Abstract

Infection control requires integrating behavioural dynamics into epidemic models. However, models often overlook behavioural complexity due to limited empirical data. We adopted an interdisciplinary approach to develop a modelling-oriented behavioural dataset.

We applied the *Capability*, *Opportunity*, *Motivation*-Behaviour (COM-B) framework to assess COVID-19 vaccination behaviour from 22,228 survey responses across six European countries (March–July 2024). We examined how willingness to vaccinate aligned with uptake and timeliness, and traced country-specific temporal changes in perceived COVID-19 severity and willingness. To capture peer-driven opinion formation, we introduced a novel indicator – discussion contacts – measuring interactions on relevant topics.

Willingness correlated with both uptake and timeliness, yet 6-18% of initially unwilling respondents ultimately received ≥2 doses. Lower willingness was associated with a 2.0-month vaccination delay. Perceived severity and willingness to vaccinate declined throughout the pandemic. Behavioural indicators systematically varied by vaccination status, particularly in the *Motivation* domain. Daily discussion contacts followed the patterns of in-person contacts, ranging from 1.4–2.9 for adults <60 years to 0.5–1.2 for those ≥60 years.

This dataset offers theory-informed and time-sensitive inputs to support the development of more realistic and policy-relevant behavioural-epidemic models.

## INTRODUCTION

In the past decades, the mutual influence between human behaviour and the spread of communicable diseases has grown increasingly evident [1]. Changes in epidemic pressure can drive adherence levels to protective behaviours during an outbreak, such as self-isolation, social distancing, mask use, hygienic measures, or vaccine uptake [2,3]. In turn, the adoption or abandonment of such behaviours can impact disease transmission and re-shape the effectiveness of infection prevention and control interventions through a dynamic feedback loop. This underscores the importance of integrating knowledge on behavioural dynamics into policy-relevant epidemic models [4,5].

Behaviour is shaped by the complex interplay between social, economic, political, and environmental factors at the individual, community, and societal levels, and it can vary significantly across countries, social groups, and over time [6]. Collecting the appropriate data to investigate such complex dynamics is a challenging task that requires a strong foundation in the behavioural and social sciences [7–9], which typically focus on who is engaging in a certain behaviour and why [5,10]. In contrast, the parametrisation of policy-relevant epidemic models requires quantitative measures of how frequently, for how long, with whom and to what extent people adopt a given behaviour [11,12]. Although behavioural complexity is broadly acknowledged [13], most epidemic models have relied on simplified behavioural representations to cope with the inherent challenges of highly nonlinear systems, often incorporating a single or very few behavioural factors at the time, such as cost-benefit [14,15] or payoff [16,17], risk perception [18], awareness [19,20] or strategic social norms [16]. While this approach has offered useful insights into selected aspects of the interaction of infection and behaviour [13], it has also contributed to the predominance of models relying on behavioural assumptions seldom grounded in established psychological, social or behavioural science theories [13,21–24]. This also prevented significant investments in targeted, interdisciplinary, theory-based and modelling-oriented data collections [11,22,23,25], resulting in few studies using empirical data on behaviour to calibrate or parametrise their epidemic models [26–28].

Even among the numerous modelling efforts prompted by the COVID-19 pandemic, only two drew on the Health Belief Model [29,30], and human behaviour was predominantly treated as an exogenous factor, incorporated through externally defined parameters [26,27]. In this approach, behavioural inputs were typically derived from survey-based data on testing adherence [31] or social contacts [32]. However, these approaches may not capture the full scope of behavioural feedback in real-world settings, leaving behaviour as a major source of uncertainty in epidemic projections [24,33].

Advancing the field of coupled behavioural-epidemic modelling thus requires the development of theory-grounded data collections that capture sufficient realism to distinguish between behavioural profiles, while enabling a parsimonious quantification of complex behavioural elements for modelling purposes [28,34,35]. To this end, we developed an interdisciplinary framework to guide the collection of empirical data on human behaviour, specifically aimed at parametrising coupled behavioural-epidemic models. We leveraged the validated “Capability, Opportunity, and Motivation” framework for behaviour change (COM-B) [36] to collect comprehensive data on population- and context-specific factors potentially associated with individuals’ vaccination behaviour in six European countries, *i.e.* Germany (DE), Spain (ES), France (FR), Hungary (HU), Italy (IT), and the United Kingdom (UK), spanning pandemic and endemic periods and different diseases, including COVID-19, influenza, and measles. In this article, we elaborate on selected aspects of the collected dataset to illustrate how this interdisciplinary approach can address the following challenges in coupled behavioural-epidemic modelling:

i. In models that are designed to predict the impact of behaviour on epidemic outcomes, psychological antecedents of behaviour, such as intention or willingness, are often employed as proxies. However, their relationship with actual behaviour is complex and continues to be an area of active investigation [37]. In this work, we empirically quantified the discrepancy between willingness to vaccinate and observed vaccination behaviour, and further analysed how varying levels of willingness influenced the timeliness of vaccination uptake.
ii. Failure to incorporate dynamic behavioural variations into data-driven epidemic models can result in oversimplified representations of real-world complexity. We analysed temporal and country-specific fluctuations in perceived COVID-19 severity and willingness to vaccinate, measured across five different time points anchored to the rollout of sequential COVID-19 vaccine doses.
iii. Complex social constructs remain inadequately represented in coupled behavioural-epidemic models. We illustrate how integrating behavioural theory and specific modelling requirements early in the data collection process can support the systematic investigation of both cognitive and social mechanisms through which behavioural choices are made, and so facilitate the definition of novel, evidence-based endogenous model components, such as peer-driven behavioural response.

## METHODS

### The interdisciplinary approach

**Fig. 1** shows the proposed interdisciplinary framework, placing the dynamic relationship between human behaviour and disease transmission at the intersection between validated behavioural science investigation and state-of-the-art epidemic modelling methodologies. Among the contemporary behavioural, psychological, and social science theories [7,38], we selected the COM-B framework, which expands the definition of behaviour beyond the traditional cognitive processes and provides guidance on how to structure comprehensive information on the potential psychological, physical, motivational, and environmental determinants of behaviour [36]. This framework has already been applied in several different settings to investigate vaccination [39–42] and other health-related behaviours [43,44], and to inform the behavioural components of epidemic models [45]. Building on this literature, we developed a comprehensive survey instrument grounded in the COM-B framework to capture the multidimensional drivers of vaccine uptake across six European countries. In parallel, we assessed how key behavioural variables could be parametrised and tailored the data collection instrument to satisfy these predefined modelling requirements. The following sections define the behavioural outcomes investigated and describe the variables captured through the survey instrument (n=91). A full version of the questionnaire is available as Online Resource 2.

**Fig. 1.**
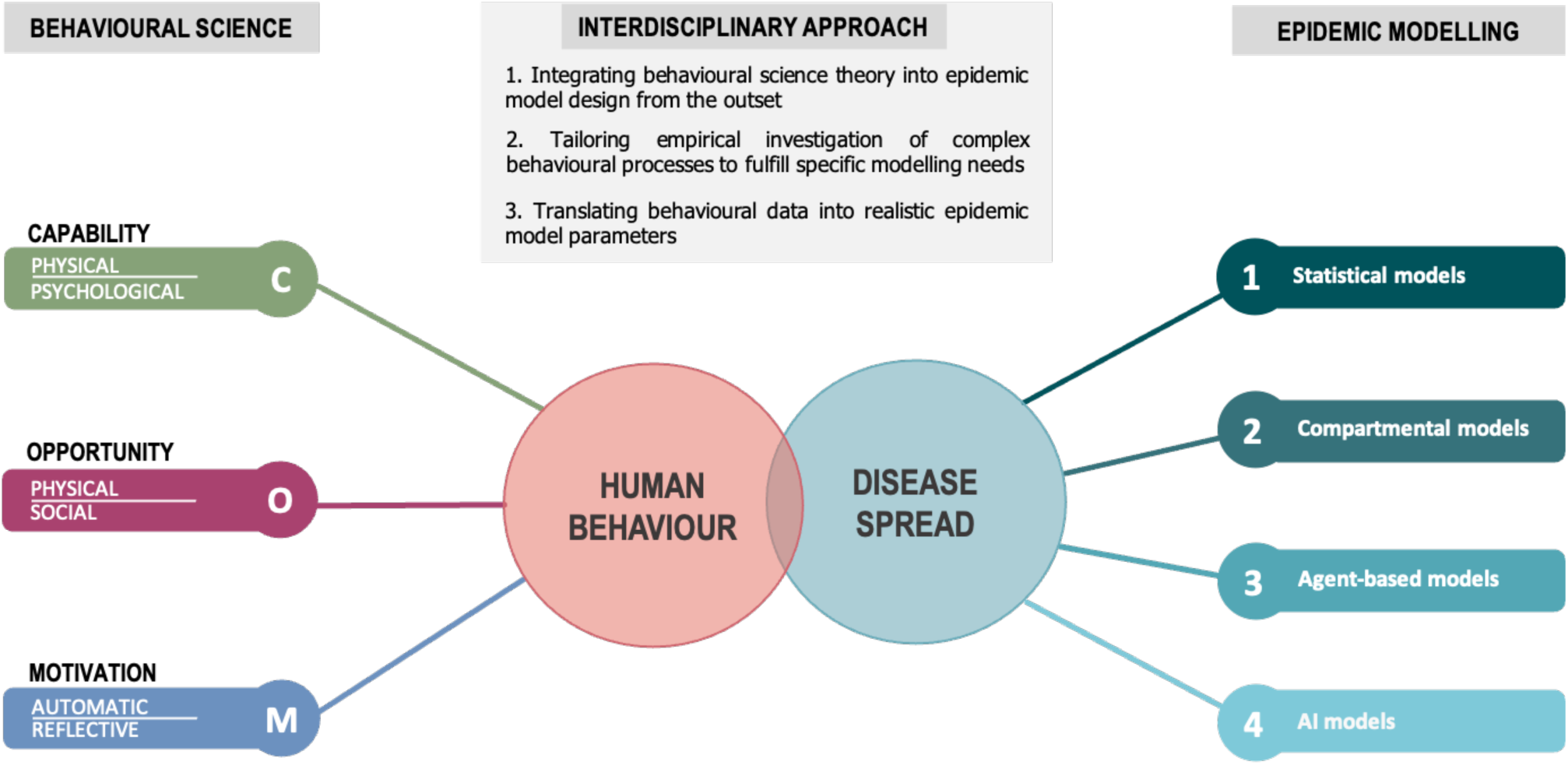
Interdisciplinary approach for the collection of empirical data on behaviour to parametrise behavioural-epidemic models. The dynamic relationship between human behaviour and disease spread lies at the core of the framework. On the left of the diagram, the “Capability, Opportunity, and Motivation” behaviour (COM-B) model dimensions [36] provide a validated structure to investigate factors potentially influencing behaviour (for more details, please see **Table 1**). On the right, state-of-the-art epidemic modelling approaches that can integrate human behaviour components endogenously.

**Table 1.**
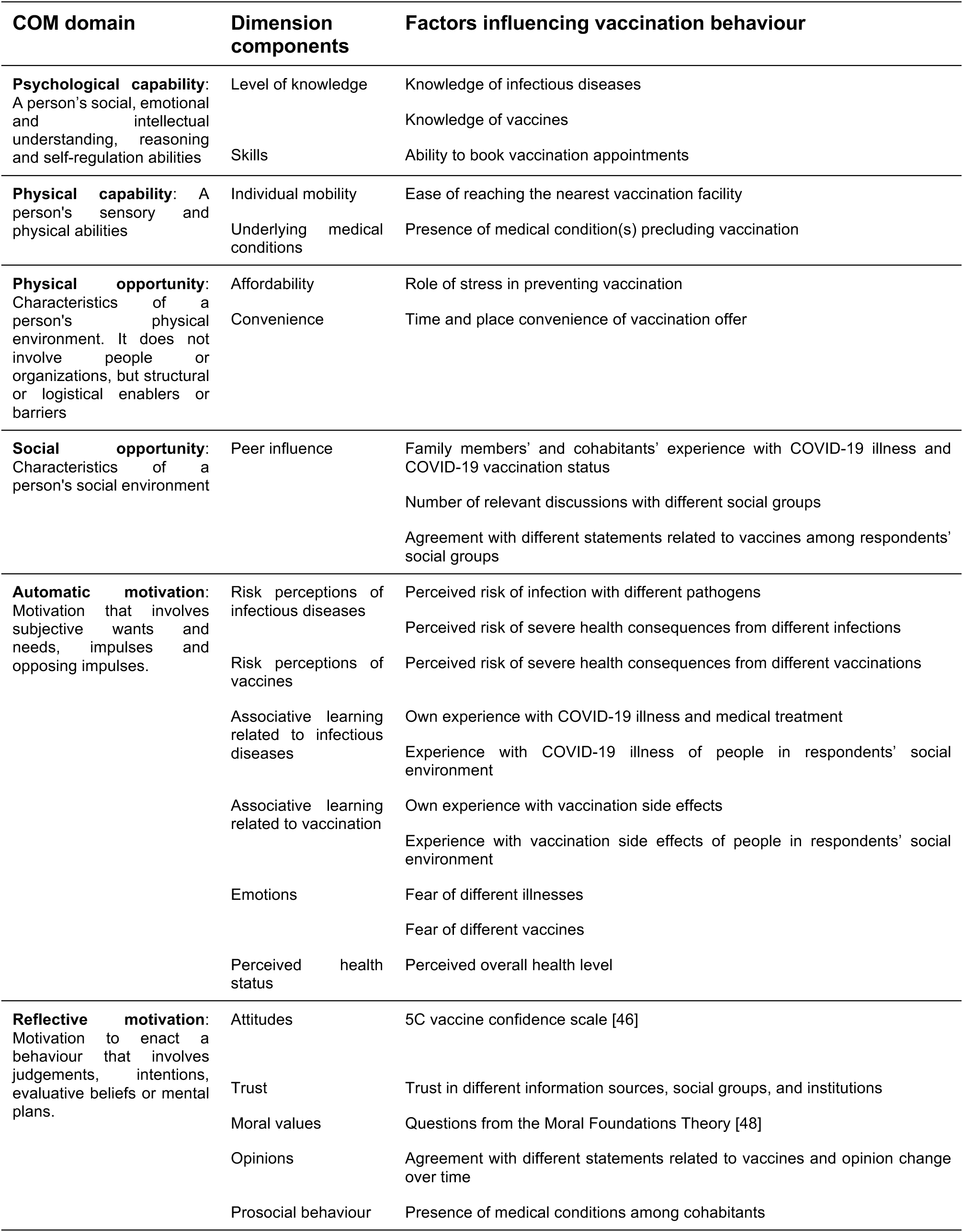
Factors potentially influencing COVID-19 vaccination behaviour, stratified by the *Capability*, *Opportunity* and *Motivation* (COM) model dimensions [36]. Factors are defined as elements that facilitate or are needed to carry out the behaviour. The survey instrument assessed each individual factor through one or more questions.

### Self-reported behavioural outcomes

The primary outcomes of the survey were the number of COVID-19 vaccine doses ever received by respondents and COVID-19 vaccination timeliness (Supplementary Material, Table S1). Respondents were also asked to report the reasons for taking up, refusing, or interrupting COVID-19 vaccination. Secondary vaccination outcomes included willingness to get vaccinated against COVID-19 over time, number of influenza vaccine doses received in the five years prior to the survey, and consent to routine childhood vaccinations for one’s child(ren), if any. In addition, we investigated respondents’ self-reported adherence to other protective behaviours at the time of the survey, *i.e.* mask use and self-isolation when experiencing respiratory symptoms or mask use in crowded places when the epidemic pressure is high (Supplementary Material, Table S1).

### Survey-based data on human behaviour for modelling

The questionnaire gathered detailed information on respondents’ socio-demographic, health, and economic characteristics, location of residence, composition of household and social groups, social identity, exposure to on- and offline information sources, and factors within the COM domains potentially associated with COVID-19 vaccination behaviour **(Table 1)**. While some questionnaire segments, such as those assessing vaccine confidence [46,47] or moral values [48], were constructed leveraging validated survey instruments, others were specifically designed to parameterise coupled behavioural-epidemic models.

Acknowledging the time-sensitive nature of “perceived risk from COVID-19” and “willingness to vaccinate against COVID-19”, we measured these retrospectively at five different time points: i) before the introduction of COVID-19 vaccines; ii) when respondents were offered the 1^st^/2^nd^ vaccine dose; iii) when respondents were offered the 3^rd^ dose; iv) when respondents were offered the 4^th^ dose; v) on the day before the survey. Only respondents who received two or more vaccine doses were asked to provide information for the third and fourth time point.

Building upon the existing concept of social contact matrices used as proxies for potential transmission events [49], we defined the number of age-specific in-person contacts as the number of different individuals with whom the respondent had a direct contact on the day prior to the survey, including an exchange of at least a few words or physical contact (i.e. a handshake, embracing, kissing, contact sports). Expanding this definition, we also introduced a new measure to proxy information diffusion events, *i.e.* “discussion contacts”, assuming these may significantly influence respondents’ opinions and subsequent behaviour. Discussion contacts were defined as the number of different individuals with whom respondents had a relevant conversation – whether in person, by phone, or online – during the week preceding the survey. Relevant conversations were those involving an exchange of information that could be useful in forming an opinion on whatever topic, from personal life choices (*i.e.* relationships, educational choices, work choices, health choices including vaccine choices) to general topics (*i.e.* current events, politics, and health, including conversations about vaccines). Both in-person and discussion contacts were stratified by social group of the contactee, *i.e.* partner, close family members, other cohabitants, friends, co-workers/classmates, or other acquaintances.

### Data collection

This study was pre-registered on Open Science Framework (OSF ID: mnb4k). The questionnaire was programmed in Qualtrics. A survey company (www.cint.com) recruited study participants from country-specific online panels in DE, ES, FR, HU, IT, and the UK, and translated the questionnaire into the five non-English study languages. The six countries were selected to capture a diverse range of political systems, cultural contexts, epidemiological conditions, healthcare infrastructures, and vaccination policies within the European context. To enable cross-country comparisons, responses related to educational levels, geographical distribution, politics, religion, and income were mapped to standardized categories (Supplementary Material, Tables S2 and S3).

In each country, the company composed a random sample of residents aged ≥18 years, selected to be representative of the national adult population based on age, sex, education level, and geographical distribution, as per 2021 EUROSTAT Statistics (https://ec.europa.eu/eurostat/databrowser/; datasets "demo_r_d2jan" and "edat_lfs_9901"; the latest available data for the UK refer to 2019). Educational levels were defined according to the International Standard Classification of Education (www.ec.europa.eu/eurostat/statistics-explained/index.php?title=International_Standard_Classification_of_Education_(ISCED)), while geographical distribution followed the Nomenclature of Territorial Units for Statistics (NUTS) level-1 classification (www.ec.europa.eu/eurostat/web/nuts). On a weekly basis during data collection, the number of respondents in each category was compared against the corresponding official population statistics. If a subgroup was underrepresented, recruitment of participants in that population was intensified.

Data collection was conducted between March 22^nd^ and July 30^th^, 2024, and concluded once ≥3,500 responses were collected per country (for details on the sample size calculation, see “Additional information” in Supplementary Material). Respondents who submitted a complete response received a pre-agreed compensation through the survey company.

### Data quality

The functionality and parsimony of the survey questionnaire were assessed by external experts in the fields of behavioural science, psychology, demography, political science, sociology, data science, infectious disease epidemiology and epidemic modelling. In addition, qualitative cognitive interviews (n=11) were conducted with members of the general public recruited through flyers distributed in public spaces. Interviewees spanned diverse age, gender, and educational characteristics. The cognitive interviews aimed to evaluate question clarity, interpretability, and relevance. Interviewees were asked to ‘think aloud’ as they answered survey items, allowing us to identify ambiguities, assess comprehension, and refine wording to ensure consistent understanding across demographic groups.

Validity and attention checks were included in the survey (see “Additional information” in Supplementary Material). To further minimise fraudulent responses, we removed respondents providing automated and duplicated responses (Supplementary Material, Table S4). After data collection, we filtered out additional records deemed unreliable according to standard criteria, and flagged inconsistencies affecting specific sections of the survey (Supplementary Material, Table S4). Flagged responses were excluded from different analysis sections only if they affected the variable(s) under study.

Post-stratification weighting adjustments were applied based on age group (18-29, 30-59, and 60+ years old), sex (Female, Male), and NUTS level-1 to correct post-data collection discrepancies with the national population (https://ec.europa.eu/eurostat/documents/203647/20621087/EU+LFS+DOI+2024.pdf). Weights were applied in the calculation of mean and percentage quantities to ensure the survey results accurately reflected the population of interest.

### Data analysis

We performed descriptive analyses to investigate willingness to vaccinate, vaccine uptake, perceived severity, relation across COM domains, and contact patterns. We computed percentages of respondents by vaccine dose number and willingness level, and by combinations of willingness and perceived COVID-19 severity over time. We defined delays in vaccine uptake as the number of months elapsed between the country-specific time point when the first dose was offered to each age group and the self-reported vaccination date for each respondent. We computed average delays and constructed bootstrap-based confidence intervals for each willingness and age group.

Respondents were categorized as having high willingness if they rated their willingness to vaccinate as "Very" or "Extremely," and low willingness if they selected "Not very" or "Not at all". Average scores on selected COM domains were derived from Likert-scale responses, which were treated as continuous variables, while distinguishing between vaccinated (at least one does of COVID-19 vaccine ever) and unvaccinated respondents. In agreement with previous work [50], the daily number of contacts was capped at 30 to limit the influence of outliers.

## RESULTS

### Study population

We collected a total of 30,429 survey responses across the six countries. After applying predefined filters for data quality (Supplementary Material, Table S4), we excluded 5,172 fraudulent and 3,029 implausible responses, retaining a total of 22,228 records for analysis (DE n = 3,890; ES n = 3,686; FR n = 3,627; HU n = 3,529; IT n = 4,196; UK n = 3,300). Among these, we flagged those with inconsistencies in COVID-19 infection or vaccination dates (n = 1,141), in-person contact numbers (n = 7,214), and discussion contact numbers (n = 8,686), as well as records with implausible school attendance on Sundays (n = 160),decimal or negative values where not applicable (n = 61). We also flagged respondents who received the Johnson&Johnson vaccine, which follows a single-dose schedule differing from the multi-dose regimens of other vaccines included in the survey (n=50). The country-specific representativeness of the study populations and respondents’ baseline characteristics are reported in Supplementary Material, Tables S2 and S3, respectively.

### Quantifying the gap between willingness to vaccinate and actual vaccine uptake

The proportion of respondents who received at least one dose of COVID-19 vaccine exceeded 70% in Germany, Spain, Italy, and the UK (range: 73.4% in Germany – 86.0% in Spain), while notably lower uptake was observed in France (64.6%) and Hungary (55.1%) (Supplementary Material, Table S5). The uptake of pandemic vaccine doses correlated positively with the uptake of influenza vaccines between 2020 and 2024 (Spearman ⍴=0.38; range: 0.30 in Hungary – 0.55 in the UK), and to a lesser extent with self-reported adherence to other protective behaviours at the time of the survey (Supplementary Material, Fig. S1).

To empirically validate psychological antecedents as proxies for actual behaviour, we charted data on respondents’ stated willingness to get vaccinated against COVID-19 when the first/second dose was offered to them and their vaccine uptake status at the time of data collection (**Fig. 2a**). In all countries, the proportion of survey respondents taking up 1, 2, 3, or 4+ COVID-19 vaccine doses increased with respondents’ initial willingness to get vaccinated (**Fig. 2a**; Supplementary Material, Table S5). The proportion of unvaccinated respondents was highest among those stating no initial willingness to get vaccinated (range: 78.2% in Spain – 89.3% in the UK), and lowest among the “extremely willing” (range: 0.4% in Italy – 2.7% in Hungary) (Supplementary Material, Table S5). Nonetheless, a non-negligible percentage of respondents in the “not at all willing” group took up ≥2 doses (range: 5.7% in the UK – 17.9% in Spain). Moreover, a notable percentage of respondents who were initially extremely willing to get vaccinated stopped vaccinating after the first or second dose (range: 7.7% in France – 18.3% in Spain).

**Fig. 2.**
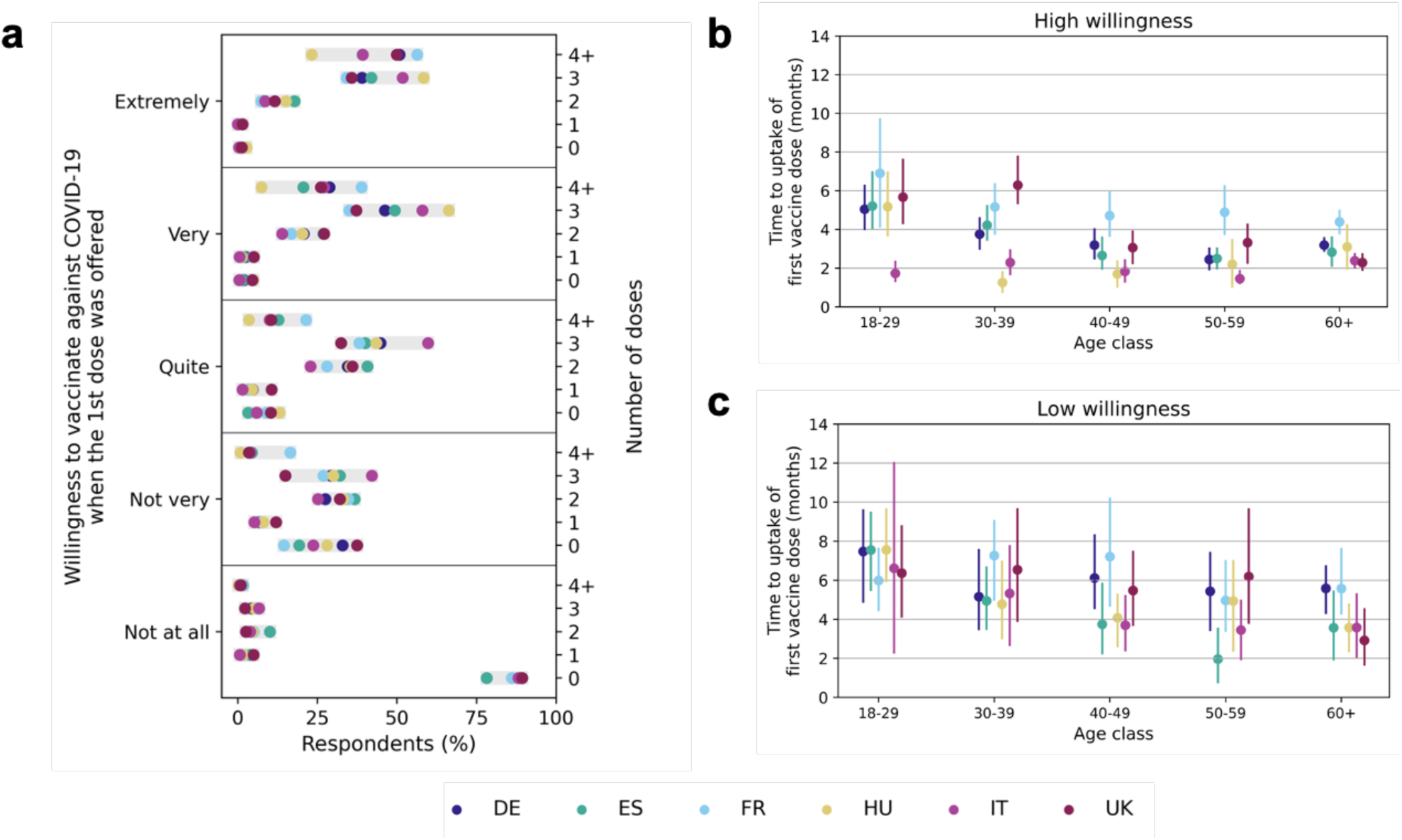
Willingness-behaviour gap and timeliness of COVID-19 vaccine uptake. **a.** Percentage of respondents taking up 0, 1, 2, 3, or 4+ doses of COVID-19 vaccine, by country and stated willingness to get vaccinated at the time the first dose was offered to them. The plot excludes respondents who: i) indicated that COVID-19 vaccination was mandatory for them (DE n=573; ES n=540; FR n=704; HU n=517; IT n=637; UK n=442); ii) indicated wanting the mandatory vaccination certificate (DE n=904; ES n=783; FR n=1,436; HU n=638; IT n=1,305; UK n=560); iii) provided free-text responses as main reasons for vaccination (DE n=164; ES n=59; FR n=42; HU n=107; IT n=102; UK n=48); iv) reported a medical condition that prevented them from getting vaccinated (DE n=15; ES n=4; FR n=12; HU n=24; IT n =15; UK n =2); v) stopped vaccinating because they experienced COVID-19 illness (DE n=526; ES n=243; FR n=479; HU n=140; IT n=646; UK n=203). Respondents who were administered 1 (n=21), 2 (n=21), or 3 (n=5) Johnson&Johnson vaccine doses were recoded as having received a total of 2, 3 or 4 doses, respectively. For more details, please refer to Supplementary Material, Table S4. **b.** Average number of months between the time point (MM/YYY) when the COVID-19 vaccine was first offered to each specific age group and when vaccinated respondents in that group received their first dose, by country. The panel includes respondents who were ‘not very’ or ‘not at all’ willing to get vaccinated at the time the first dose was offered to them. Vertical bars indicate bootstrap-based confidence intervals. **c.** As in **b** for those who indicated they were ‘very’ or ‘extremely’ willing to get vaccinated. For delay, reference dates were primarily sourced from the Oxford dataset, which includes information on vaccination openings or national coverage registers [51]. For more details, please refer to Supplementary Material, Table S5. In both panels **b** and **c**, the weights reflect 10-year age groups, as shown in the plots, along with sex. Both panels **b** and **c** exclude respondents who: reported i); ii); iii) as in **a**; iv) indicated uptake dates preceding the vaccine offer to their age category (DE n=574; ES n=835; FR n=447; HU n=430; IT n=901; UK n=683); v) were administered the Johnson&Johnson vaccine, which was offered to the population with a delay (DE n=11; ES n=8; FR n=6; HU n=17; IT n=6; UK n=2).

We investigated age-specific vaccination timeliness according to respondents’ initial willingness to vaccinate (**Fig. 2b and 2c**; Supplementary Material, Table S6). On average, respondents aged 18-29 years took 5.6 months to receive their first dose after the vaccine was made available to their age group (delay range: 2.8 months in Italy – 6.6 months in Hungary). In comparison, elderly aged 60+ years received the vaccine on average within 3.3 months of its availability to their age group (delay range: 2.5 months in the UK – 4.7 months in France).

In all countries, vaccination timeliness increased with initial willingness to get vaccinated. Those with high initial willingness received their first dose on average within 3.0 months (delay range: 2.0 months in Italy – 5.0 months in France) (**Fig. 2b**; Supplementary Material, Table S6). In comparison, those with low initial willingness delayed their first dose by 2.0 additional months on average (delay range: 4.1 months in Italy – 6.2 months in France) (**Fig. 2c**; Supplementary Material, Table S6).

### Temporal and country-specific variations of behavioural factors

We investigated how two potential behavioural determinants, *i.e.* perceived COVID-19 severity and willingness to take up the COVID-19 vaccine, changed over time and across countries. These two variables were retrospectively measured at five different time points. Before the vaccine was offered (**Fig. 3**, first column), the percentage of respondents reporting high or extremely high levels of perceived COVID-19 severity ranged from 32.4% in Germany to 44.9 % in Italy. Willingness to vaccinate against COVID-19 showed a greater variation across countries, with percentages of respondents reporting high or extreme willingness to vaccinate against COVID-19 ranging from 27.0% in Hungary to 61.7% in the UK. Referring to the same point in time, a considerable share of respondents reported they were not at all willing to get vaccinated both in France (39.1%) and Hungary (42.5%), while lower values were recorded in all other countries (range: 13.0% in the UK - 19.4% in Germany).

**Fig. 3.**
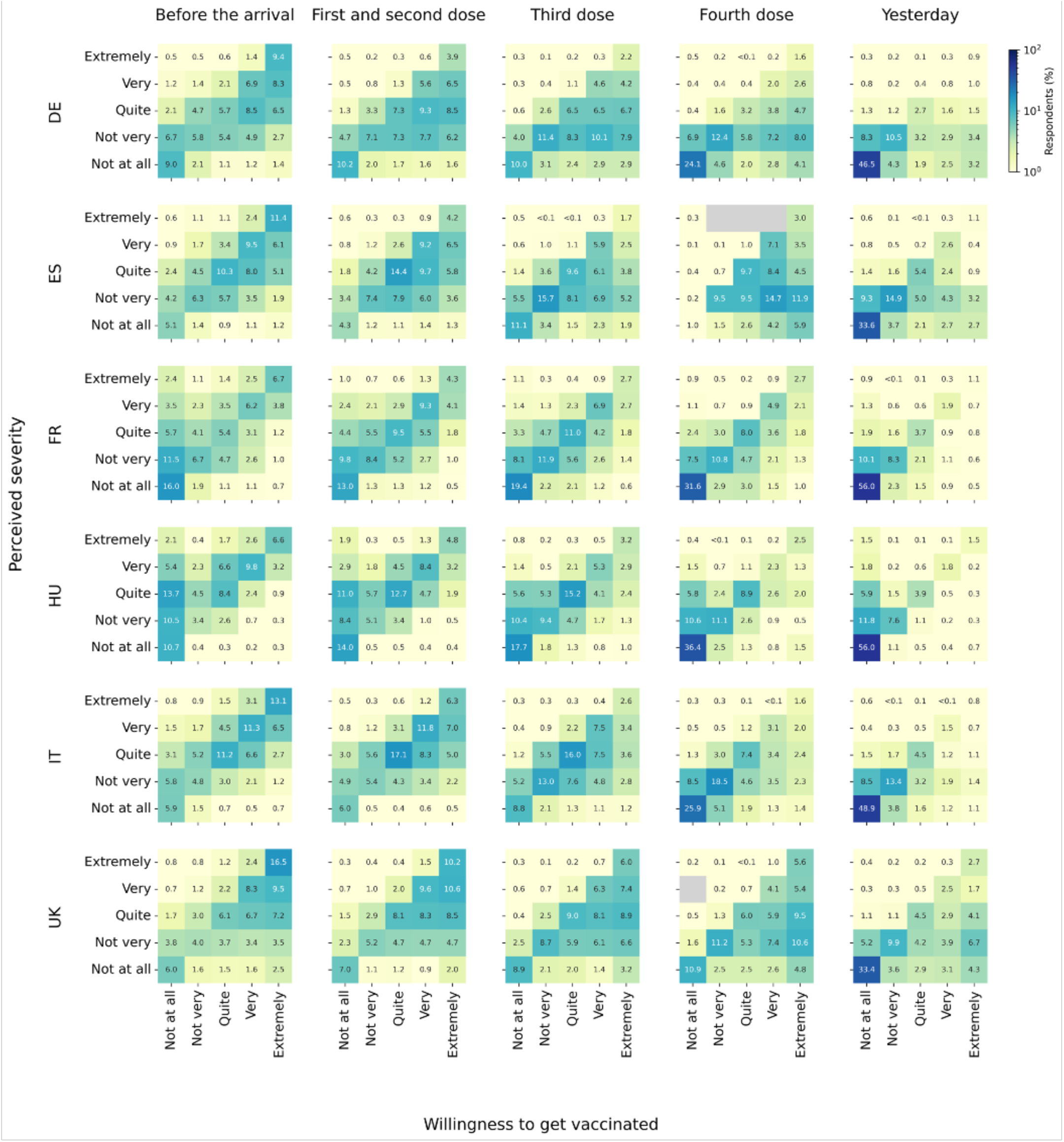
Temporal and country-specific variation of behavioural factors. Percentage of respondents indicating different levels of perceived COVID-19 severity and willingness to get vaccinated against COVID-19 over time and by country. Timepoints: t1 = Before the introduction of the COVID-19 vaccine; t2, t3 and t4 = When the 1^st^ and 2^nd^, 3^rd^, or 4^th^ vaccine dose were offered to respondents, respectively; t5 = the day before the survey. Only respondents who received two or more vaccine doses were asked to provide information for t3 and t4. Grey shading indicates that no respondents were in that specific group.

When considering the time when the first vaccine dose was offered to them (**Fig. 3**, second column), the share of respondents expressing high or extremely high perceived COVID-19 severity had decreased in all countries (range: 20.2% in Germany – 36.8% in the UK), while high or extremely high willingness to vaccinate remained stable in all populations (range: 26.7% in Hungary – 61.0% in the UK). Among respondents who received two or more doses, perceived severity and willingness to get additional COVID-19 vaccine doses shifted towards lower levels as the vaccination campaign progressed (**Fig. 3**, third and fourth column). When considering the most recent data point, *i.e.* the day before the survey (**Fig. 3**, last column), larger proportions of respondents reported low or no perceived COVID-19 severity (range: 77.3% in the UK – 86.9% in DE), and the majority indicated no or low willingness to get vaccinated against the disease (range: 55.5% in the UK – 87.6% in Hungary).

### Disentangling behavioural mechanisms across COM-B dimensions

Grounding the empirical data collection in a validated behavioural model allowed us to collect abundant information on potential behavioural determinants, and to systematically assess complex mechanisms underlying behavioural choices. **Fig. 4** shows respondents’ average responses to a selection of survey questions, reflecting different elements of the *Capabilty*, *Opportunity*, and *Motivation* components of behaviour (Supplementary Material, Table S7). Responses are grouped by respondents’ COVID-19 vaccination status, with “vaccinated” respondents including anyone who received at least one dose ever.

**Fig. 4.**
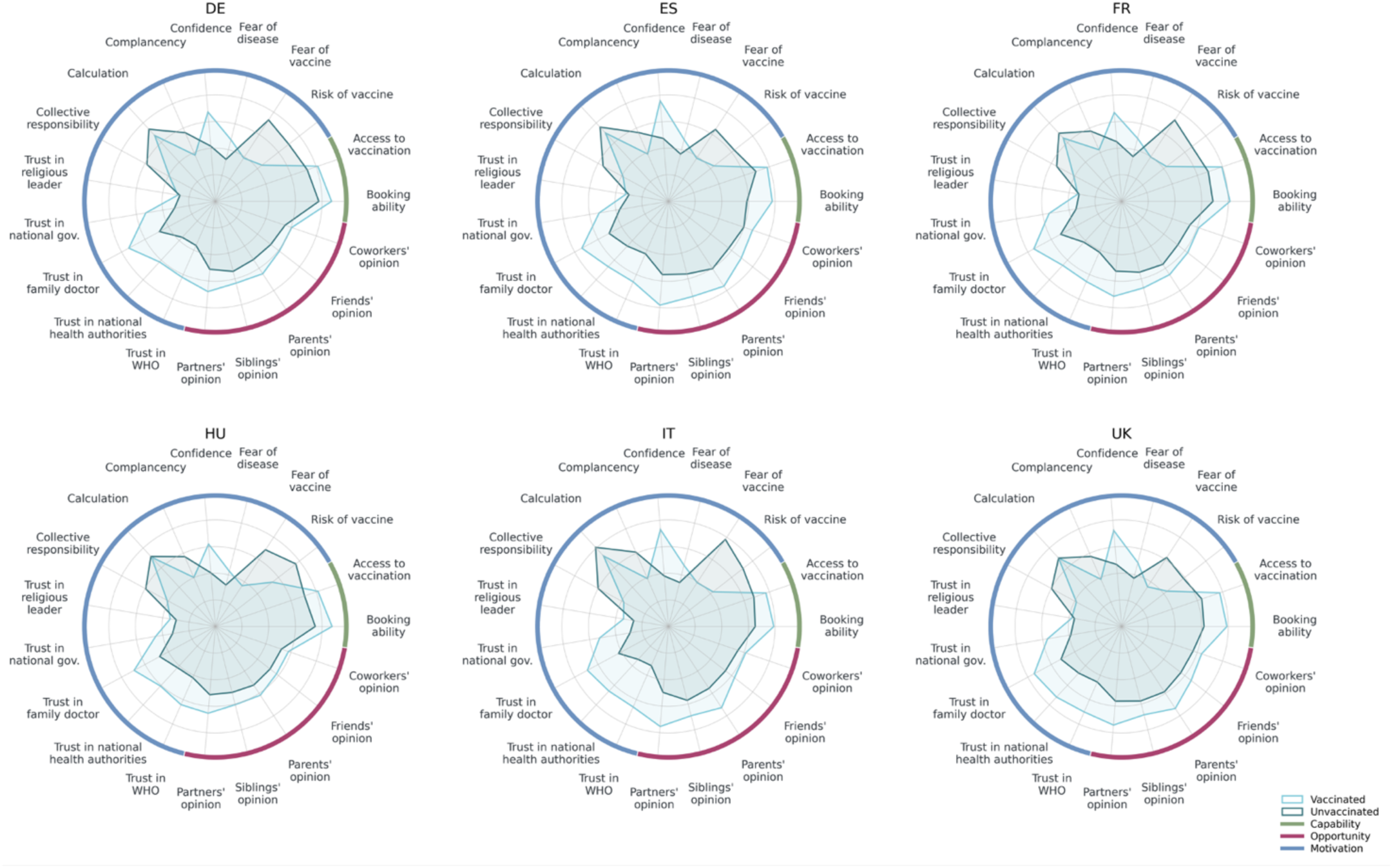
Systematic mapping of factors potentially associated with COVID-19 vaccination behaviour. Radar charts with weighted means of Likert scale values chosen by respondents in response to selected survey questions across COM-B model dimensions, stratified by country and respondents’ COVID-19 vaccination status. Vaccinated respondents include those who took up at least one dose of the COVID-19 vaccine ever. In the Opportunity section, responses only include those from respondents who previously reported the presence of each given social group. For this variable, average values show each social group’s level of agreement with the view that vaccines are essential for protecting human health, as reported by respondents. The plot excludes respondents who i) indicated that COVID-19 vaccination was mandatory for them (DE n=573; ES n=540; FR n=704; HU n=517; IT n=637; UK n=442); ii) indicated wanting the mandatory vaccination certificate (DE n=904; ES n=783; FR n=1,436; HU n=638; IT n=1,305; UK n=560); iii) provided free-text responses as main reasons for vaccination (DE n=164; ES n=59; FR n=42; HU n=107; IT n=102; UK n=48); iv) reported a medical condition that prevented them from getting vaccinated (DE n=15; ES n=4; FR n=12; HU n=24; IT n =15; UK n =2). Questionnaire items corresponding to the plotted variables are listed in Supplementary Material, Table S7. Respondents answered these questions by selecting a value on 5-point or 7-point scales. For the radar plots, 7-point scales were transformed into 5-point scales by pooling the three central options (“Slightly disagree”, “Neutral”, and “Slightly agree”). Average values are reported in Supplementary Material, Table S8.

Average responses to most of these questions varied according to COVID-19 vaccination status, and the observed patterns were qualitatively very similar across all countries (**Fig. 4**; Supplementary Material, Table S8). The most prominent differences were observed in the *Motivation* dimension. On average, unvaccinated respondents expressed higher fear of the COVID-19 vaccine (range unvaccinated: 3.1-3.9; range vaccinated: 1.8–2.0) and higher perceived likelihood to experience severe health consequences from vaccination (range unvaccinated: 2.9-3.8; range vaccinated: 2.1-2.7), but lower fear of COVID-19 disease (range unvaccinated: 1.6-1.9; range vaccinated: 2.1-2.5). Vaccine confidence was also consistently lower among the unvaccinated across all study populations (range unvaccinated: 1.9-2.4; range vaccinated: 3.1-3.8). In every country, mean levels of trust in both the national government and different public health authorities were always lower in the unvaccinated group.

In the *Opportunity* domain, unvaccinated respondents across countries reported stronger disagreement within their close social groups with the view that vaccines are essential for protecting human health (range unvaccinated: 2.5–3.0; range vaccinated: 2.9–3.9).

In the *Capability* dimension, perceived ability to book vaccination appointments (range unvaccinated: 2.9-3.9; range vaccinated: 3.9-4.4) and ability to reach the nearest vaccination facility (range unvaccinated: 3.2-3.6; range vaccinated: 3.9-4.1) were on average lower among unvaccinated respondents in all countries.

Similar, albeit smaller, differences were observed between respondents who received one or more influenza vaccine doses in the previous five years and those who never received one, again most visible in the *Motivation* dimension (Supplementary Material, Fig. S2; Table S8).

### Introducing new behavioural dimensions for coupled behavioural-epidemic modelling

To enable the investigation of how peer communication may shape individual opinions and subsequent behaviours, we amplified the well-established quantification of in-person social contacts for infectious disease modelling [49] by complementing it with the new dimension of discussion contacts (**Fig. 5**; Supplementary Material, Table S9). Across all countries, the elderly aged 60+ consistently reported the lowest average number of daily discussion contacts, ranging from 0.5 in Spain to 1.2 in the UK (**Fig. 5a**; Supplementary Material, Table S9). Among those under 60, contact patterns varied by country. In Germany, discussion contacts remained relatively stable across age groups, with averages ranging from 1.4 among 18–29-year-olds to 1.7 among those aged 40–49 years. Similarly, in the UK the number of discussion contacts was on average 2.5 for age groups up to the 40–49-year-olds and decreased to 1.4 among those aged 50–59. In contrast, a progressive decline with age was observed in Spain, where contact numbers dropped from 2.8 in those aged 18–29 to 2.0 in the 50-59 age group. A distinct pattern emerged in France, Hungary, and Italy, where the average number of discussion contacts peaked among those aged 30–39 years, reaching 2.4 in France, 2.3 in Hungary, and 2.9 in Italy.

**Fig. 5.**
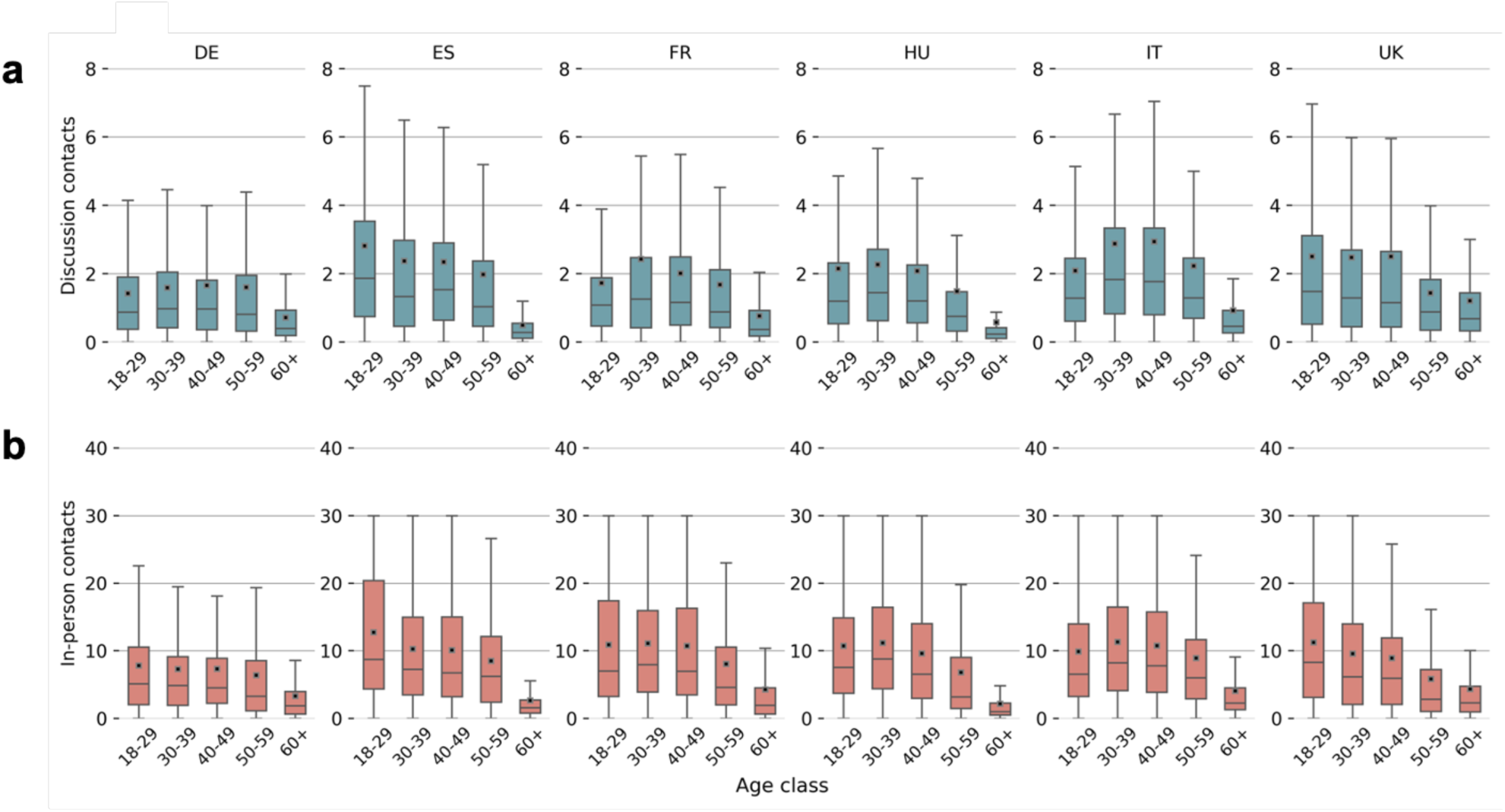
A new dimension of social contacts. **a.** Number of daily discussion contacts by age group and country. The number of weekly discussion contacts as assessed in the survey was recalculated as daily. Boxes represent medians and interquartile ranges. Whiskers represent minimum and maximum values capped at 30 daily contacts, with values >30 being imposed to 30. Small rectangles represent average values. **b.** As in a for in-person contacts by age group and country. In both panels, respondents flagged as reporting implausible contact numbers were excluded (DE n=1,452; ES n=1,725; FR n=1,395; HU n=1,561; IT n=1,780; UK n=1,180).

The average number of daily discussion contacts was strongly correlated with in-person contacts (Pearson r= 0.93) (Supplementary Material, Fig. S3). In all countries, individuals aged 60+ years reported the lowest number of in-person contacts, ranging from 2.2 in Hungary to 4.4 in the UK (**Fig. 5b**; Supplementary Material, Table S9). In Hungary and Italy, the highest levels of in-person contacts were observed among adults aged 30–39 years with 11 daily average contacts. In contrast, the number of in-person contacts peaked at 11.3 and 12.7 among younger adults aged 18–29 years in the UK (11.3) and Spain, respectively. A similar average number of daily in-person contacts was reported in all age groups <50 years in Germany (range: 7.3-7.8). Distinguishing by vaccinated and unvaccinated individuals, no substantial differences on either average daily discussion or in-person contacts were observed at the time of data collection (Supplementary Material, Fig. S4).

## DISCUSSION

In response to growing calls for greater collaboration across scientific disciplines [12,24,28], we implemented a novel interdisciplinary approach to address a longstanding challenge: integrating the dynamic nature of human behaviour into epidemic models. Acknowledging the limitations of traditional behavioural science methods in generating data that can be readily incorporated into epidemic models [11,12], we systematically integrated expertise from mathematical modelling, epidemiology, and behavioural and social science to design and implement a large-scale, nationally representative survey across six European countries. This unique dataset was developed with the explicit goal of supporting the construction of model components that reflect the multifaceted, context-dependent nature of vaccine intentions, uptake, and delay, thereby enhancing the realism and applicability of coupled behavioural-epidemic simulations.

By quantifying the difference between willingness to vaccinate and actual vaccine uptake, we provided empirical evidence to refine the use of psychological antecedents as proxies for actual behaviour to inform epidemic scenarios. Our findings are aligned with the psychological literature [52–55], indicating that intentions alone rarely provide a complete explanation for human behaviour [37,55]. This was also corroborated by survey findings during the COVID-19 pandemic, where both vaccine uptake [56,57] and timeliness [58,59] were mediated by vaccine hesitancy despite respondents’ positive intentions. Accordingly, the uptake of sequential vaccine doses was negatively affected by adverse experiences with the first dose [60] and growing vaccination fatigue [61,62].

These behavioural nuances have critical implications on policy planning and effectiveness, because models relying on psychological antecedents alone may misestimate the timing, magnitude, or trajectory of epidemic waves. Beyond the specific concerns associated with the intention-behaviour gap, the absence of a validated theoretical foundation presents further challenges for coupled behavioural-epidemic modelling. For instance, it may hinder the accurate differentiation of related behavioural constructs, such as risk perception and fear, which are often perceived as conceptually similar. Specific behavioural constructs may also be presumed to have a direct influence on behavioural choices, despite a lack of consistent evidence supporting this. A plethora of theories and conceptual approaches dedicated to investigating the multifaceted nuances of human behaviour are available, and published reviews can help navigate it [7]. We found the COM-B framework model to be well-suited for our purpose, as it offered a clear, evidence-based structure, while retaining sufficient flexibility to adapt the survey tool to our specific research questions and modelling objectives.

By reconstructing the distribution of respondents according to different degrees of perceived COVID-19 severity and vaccine willingness during critical phases of the vaccine rollout, we uncovered considerable temporal and country-specific variations. Fluctuations in perceived COVID-19 severity and willingness to vaccinate were also frequently observed in other survey initiatives conducted across Europe either concurrently or prior to our study [63–65], with hesitant individuals exhibiting the greatest variability over time [65]. Previous studies showed that such variations were driven by infection rates [66,67], socio-demographic attributes [68], vaccination status and media exposure [67,69,70]. Our dataset offered the opportunity to examine the relationship between these two variables using validated behavioural scales within the same sample. Such high-resolution empirical data that capture spatio-temporal behavioural variations can significantly improve model structures and reduce assumptions that fail to account for changes in public responses to health threats [71,72].

Our dataset provided a solid foundation to disentangle behavioural mechanisms across multiple behavioural dimensions. Using the COM-B model, we identified distinct behavioural profiles associated with vaccination status. Key differentiators included fear of side effects, perceived risk, vaccine confidence, and trust in family doctors. These results align with findings from previous surveys conducted in Europe. Fear of side effects and concerns about the rapid vaccine development were the most commonly reported reasons for vaccine refusal or delay in Italy [58], while low perceived risk of COVID-19 and distrust in health authorities were key drivers of low uptake in France and Germany [73,74]. In France, vaccinated respondents exhibited higher confidence in vaccine safety, likely due to selective information exposure [75]. Trust in family doctors and experts also emerged as a strong positive predictor of uptake in this country [73], consistent with previous findings from Hong Kong and Singapore [74]. Differences in the *Opportunity* dimension appeared to be less pronounced between vaccinated and unvaccinated individuals, both in terms of average opinion towards vaccination in selected social groups and average number of discussion contacts. This could suggest that behavioural homophily - while present - may not be the dominant driver of observed differences across vaccination statuses.

In our study population, vaccine uptake also correlated with other behaviours. This is in line with UK data from 2021, where respondents who received two doses of the COVID-19 vaccine were more likely to resume social activities, and those with greater motivation, opportunity, and capability continued engaging in preventive measures such as mask-wearing and distancing [76].

Together, these findings underscore the importance of examining the full range of behavioural domains in an integrated manner rather than in isolation, as varying combinations of social and cognitive factors may lead to distinct patterns of compliance to protective measures. The overall effect of the epidemic response may depend on the interconnection of individual adherence decisions [77]. Nevertheless, most epidemic models reduce behaviour to in-person contact data, overlooking the complexities of behavioural elements, and particularly neglecting those related to complex social interactions, such as the peer-driven formation and diffusion of opinions [29,30,45,78]. In our study population, the distribution of discussion contacts closely mirrored that of in-person contacts in the same country, suggesting that discussion interactions may represent a small-scale projection of broader social interactions. By embedding mechanisms such as contact heterogeneity, peer influence, and social learning – and by accounting for how these social interactions affect disease spread – coupled behavioural-epidemic models will better reflect the complexity of real-world dynamics [79].

Several limitations must be acknowledged. First, the retrospective self-reported nature of behavioural and cognitive data – particularly across multiple time points – may introduce recall bias. To mitigate this risk, we employed memory aids, such as visual timelines, and anchored questions to respondents’ lived experiences to help them identify relevant time frames. For instance, perceived COVID-19 severity was assessed for time points chosen to correspond to key milestones of each country’s vaccination campaign. Despite its limitations, the retrospective design enabled analyses across time and contexts, including complex constructs such as the intention-behaviour gap - otherwise requiring resource-intensive longitudinal studies. Second, the use of online survey panels may pose risks of selection bias, impacting both the representativeness of the sample and the validity of the responses. To address this concern, nationally representative quotas were established based on key demographic variables, and recruitment was adjusted in real time to ensure these quotas were achieved. Comparison of vaccine coverage and socio-demographic variables with national statistics and similar surveys [80] did not reveal major recall or selection biases. Third, participants under the age of 18 were excluded from this study, as they were not considered the primary decision-makers for vaccination or related behaviours. However, respondents who were parents of at least one child under 10 years of age were asked to report their child(ren)’s vaccination status for both COVID-19 and routine childhood immunisations, as well as the reasons underlying their acceptance or refusal of COVID-19 vaccination for their child(ren). Finally, although the generalisability of the study findings beyond Europe or the COVID-19 context may be limited without cultural and epidemiological adaptations, the underlying methodological approach presented here can be extended to any setting, time period, behaviour, or disease.

### Conclusions

Despite the recent surge in coupled behavioural-epidemic modelling studies prompted by the influenza and COVID-19 pandemics, truly integrative advances remain limited [26–28]. This work provides a robust empirical foundation for a new generation of parsimonious yet realistic models that better reflect behavioural complexity, improve disease trend forecasting, and strengthen policy relevance. Our theory-informed design supports both rigour and reproducibility, extending past efforts [81] to systematise behavioural integration into epidemic models.

Emerging approaches that incorporate Artificial Intelligence (AI) into epidemic modelling [82,83] further underscore the need for rich, high-resolution behavioural datasets capable of training large language models to simulate human decision-making and behavioural dynamics. Though cross-sectional by design, our dataset offers a robust combination of behavioural depth, theoretical coherence, and population-level representativeness, positioning itself as a valuable resource for advancing both conventional and AI-driven behavioural-epidemic models. Looking ahead, longitudinal frameworks for collecting [84,85] and sharing (www.ccp.jhu.edu/kap-covid/) behavioural data would substantially increase their applicability in coupled behavioural-epidemic models. In health crisiscontexts, where time and resources are constrained, pre-established data systems become essential [35,86,87]. Embedding behavioural modules into participatory surveillance platforms offers a promising solution (www.influweb.org/about; www.influenzanet.info/home).

Equally important is the integration of different data sources. Triangulating survey responses with observational data from separate sources can help assess their reliability and mitigate systematic biases [84,88,89]. Given the increasingly intertwined nature of human behaviour across on- and offline environments, emerging data streams, such as those from social media or personal digital devices, offer valuable opportunities to capture high-resolution, passively collected behavioural indicators [2,90]. For instance, a number of respondents in our survey provided access to their social media content. The publicly available information on their profiles, such as the content of posts, comments, and messages shared publicly or on open groups, websites followed, or "likes”, will allow us to connect and cross-validate information from both the physical and digital realms.

Ultimately, advancing the coupled behavioural-epidemic modelling agenda will demand sustained interdisciplinary collaboration, supported by the development of a shared conceptual and methodological language, as well as a high degree of flexibility and compromise. Encouragingly, promising inter-disciplinary initiatives are already emerging at the national [91,92] and international level (www.who.int/teams/one-health-initiative; www.prevention.ecdc.europa.eu/public-home).

Bridging disciplinary divides to incorporate behavioural complexities into epidemic modelling is no longer optional, it is a necessary step towards the development of more responsive, empirically grounded, and resilient public health systems.

## Acknowledgments

We acknowledge precious input for both the conceptualisation of the scientific approach and the development of the survey tool from Delia Baldassarri, Alain Barrat, Chris Bauch, Simon Cauchemez, Laura Di Domenico, Anthony Hauser, Sune Lehmann, Anna Sára Ligeti, Susan Michie, Saad Omer, Daniela Paolotti, Nicola Perra, Daniela Perrotta, Piero Poletti, Renu Singh, Ádám Stefkovics, Emilio Zagheni, Alfonso de Miguel and Carlos Gracia. We thank researchers from the Dondena Centre at Bocconi University for providing feedback on the survey tool. We thank the Dondena Centre for hosting the research project IMMUNE and for providing continuous support.

## Author contributions

AM conceived the study and led the research project. AM, MC, LL, LPL, and VO conceptualised and developed the questionnaire, with inputs from CC and DB. FT, JK, MK, PM, VC, AA and YM provided domain expertise throughout study design, data collection, and quality control. LL and LPL led the implementation of the survey. LL, LPL, and CC ensured data quality. LL, LPL, CC, EDA and FB conducted data cleaning. EC conducted the analysis and produced the visualizations. AM, VO and EC conceptualised and wrote the manuscript. All authors interpreted the results, reviewed, and approved the final version of the manuscript.

## Ethics approval

This study was approved by the Ethics Review Board of the Bocconi University in Milan on December 23^rd^, 2022 for Italy, France, and Germany (Protocol Nr. FA000544). Two amendments to the original proposal were subsequently approved to include cognitive interviews to the questionnaire validation procedures (Protocol Nr. FA000544-01; approval on June 21^st^, 2023) and to add Spain, Hungary, and the UK as additional study locations (Protocol Nr. EA000682; approval on October 30^th^, 2023).

## Consent to participate

Consent to participation was collected through an online form at the start of the survey. All survey questions were mandatory, but “I do not know” and “I prefer not to answer” options were provided for sensitive questions, *i.e.* those related to economic status or personal health. Respondents could withdraw at any time.

## Consent to publish

This manuscript does not contain any individual person’s data in any form (including any individual details, images or videos).

## Data availability

The data that support the findings of this study will be made available upon reasonable request for scientific research purposes. For further information or to request access, please contact the corresponding author.

## Code availability

The Python code used for this analysis will be deposited in Open Science Framework (OSF ID: mnb4k) upon publication.

