## Supplementary Material for "Advancing coupled behavioural-epidemic models: An interdisciplinary framework for the collection of empirical data"

### Supplementary Tables

#### **Table S1** Behavioural outcomes assessed in this study

| **Outcome** | | **Questionnaire item** | **Response options** |
|---|---|---|---|
| *COVID-19 vaccination* | | | |
| | # of vaccine doses taken | How many doses of COVID-19 vaccine have you received? | • 1<br>• 2<br>• 3<br>• 4 or more |
| | Vaccination date | We now ask you to make the effort to specify the month and year in which you received each dose of COVID-19 vaccine. As an aide, you can find the main dates of the COVID-19 vaccine schedule in the figure below. Also remember that:<br>• the second vaccine dose was normally administered between 2 and 6 weeks after the first dose;<br>• for the Johnson & Johnson vaccine, there was no second dose. | • 1st dose MM/YYYY<br>• 2nd dose MM/YYYY<br>• 3rd dose MM/YYYY<br>• 4th dose MM/YYYY |
| | Willingness to get vaccinated over time | We will now ask you to answer some questions related to the COVID-19 vaccine at specific times. How willing were you to be vaccinated against COVID-19?<br>• Before the arrival of the COVID-19 vaccine<br>• When you were offered the FIRST and SECOND dose of vaccine<br>• When you were offered the THIRD dose of vaccine<br>• When you were offered the FOURTH dose of vaccine<br>• Yesterday | • Not at all<br>• Not very<br>• Quite<br>• Very<br>• Extremely |
| *Other vaccinations* | | | |
| | Influenza vaccine | Did you get vaccinated against flu at least once in the following time periods?<br>• Before the pandemic<br>• Autumn/winter 2020-2021<br>• Autumn/winter 2021-2022<br>• Autumn/winter 2022-2023 | • Yes<br>• No<br>• I don't remember |

| | | | |
|---|---|---|---|
| | | • Autumn/winter 2023-2024 | |
| | Routine childhood vaccinations | For your minor child/children, indicate whether you have consented to the recommended childhood vaccinations. | • Yes, to all<br>• Yes, but just to some<br>• I don't know/I don't remember |
| *Protective behaviours at the time of the survey* | | | |
| | Mask use | Please evaluate how much you disagree or agree with the following statements:<br>• I use masks in crowded places when experiencing respiratory symptoms<br>• When the epidemic pressure is high, I use masks in crowded places | • Strongly disagree<br>• Moderately disagree<br>• Slightly disagree<br>• Neutral<br>• Slightly agree<br>• Moderately agree<br>• Strongly agree |
| | Self-isolation | Please evaluate how much you disagree or agree with the following statements:<br>• I stay at home when experiencing respiratory symptoms | • Strongly disagree<br>• Moderately disagree<br>• Slightly disagree<br>• Neutral<br>• Slightly agree<br>• Moderately agree<br>• Strongly agree |

**Table S2** Representativeness of country-specific study samples For each country, the left and right columns show percentages within the study sample and in the underlying country population as per Eurostat Labour Force Survey 2022 statistics[1], respectively; the latest available data for the UK refer to 2019 data and consider only the adult population below 70 years of age. To enable cross-country comparisons, the geographical distribution followed the *Nomenclature of Territorial Units for Statistics* classification (NUTS-1)[2], and educational levels were defined to map into the *International Standard Classification of Education*[3], with low, medium and high levels corresponding to ED 0-2, 3-4, and 5-8, respectively (except for the UK).

| | | DE | | ES | | FR | | HU | | IT | | UK | |
|---|---|---|---|---|---|---|---|---|---|---|---|---|---|
| | | **Sample** | **Pop** | **Sample** | **Pop** | **Sample** | **Pop** | **Sample** | **Pop** | **Sample** | **Pop** | **Sample** | **Pop** |
| *Age group* | | | | | | | | | | | | | |
| | 18-29 | 16.1 | 15.7 | 22.7 | 15.2 | 18.7 | 17.2 | 23.5 | 16.2 | 14.6 | 14.3 | 22.2 | 18.9 |
| | 30-59 | 53.3 | 49.2 | 62.8 | 53.0 | 55.3 | 48.6 | 62.5 | 51.9 | 61.1 | 49.4 | 51.1 | 50.7 |
| | 60+ | 30.6 | 35.1 | 14.5 | 31.8 | 26.0 | 34.2 | 14.0 | 31.9 | 24.3 | 36.3 | 26.7 | 30.4 |
| *Sex* | | | | | | | | | | | | | |
| | Male | 50.4 | 49.0 | 48.1 | 48.5 | 45.4 | 47.7 | 45.3 | 47.2 | 49.1 | 48.2 | 49.8 | 48.9 |
| | Female | 49.6 | 51.0 | 51.9 | 51.5 | 54.6 | 52.3 | 54.7 | 52.8 | 50.9 | 51.8 | 50.2 | 51.1 |
| *Education level* | | | | | | | | | | | | | |
| | Low | 27.8 | 20.3 | 20.3 | 43.4 | 7.1 | 23.3 | 22.3 | 18.3 | 22.1 | 46.2 | 21.9 | 21.9 |
| | Medium | 42.9 | 51.4 | 29.9 | 23.1 | 49.2 | 42.3 | 44.6 | 56.6 | 51.5 | 38.1 | 34.5 | 36.9 |
| | High | 29.3 | 28.3 | 49.8 | 33.5 | 43.7 | 34.4 | 33.1 | 25.1 | 26.4 | 15.7 | 43.6 | 41.2 |
| *NUTS-1* | | | | | | | | | | | | | |

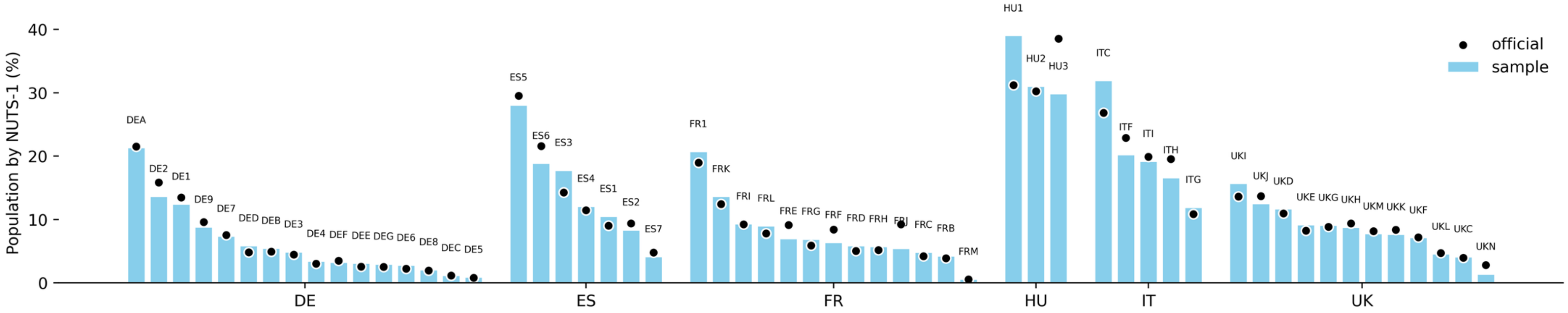

**Table S1** Participants' baseline characteristics by country In each sample, the left and right columns show percentages (%) and absolute numbers (n), respectively. This table excludes age, sex, education and NUTS-1, which are reported in Table S2. To enable cross-country comparisons, responses related to politics, income, and religion were mapped to standardized categories, *i.e.* 9 European Parliament political groups, 5 income quintiles, and 10 religious groups.

| | DE | | ES | | FR | | HU | | IT | | UK | | All countries | |
|---|---|---|---|---|---|---|---|---|---|---|---|---|---|---|
| | % | n | % | n | % | n | % | n | % | n | % | n | % | n |
| **Socio-Demographic** | | | | | | | | | | | | | | |
| *Gender* | | | | | | | | | | | | | | |
| Man | 50.5 | 1,963 | 48.1 | 1,771 | 44.6 | 1,619 | 45.3 | 1,601 | 49.1 | 2,059 | 49.5 | 1,635 | 47.9 | 10,648 |
| Woman | 49.1 | 1,911 | 51.7 | 1,905 | 55.1 | 1,997 | 54.0 | 1,904 | 50.7 | 2,128 | 50.0 | 1,649 | 51.7 | 11,494 |
| Other | 0.4 | 14 | 0.2 | 9 | 0.2 | 8 | 0.2 | 7 | 0.1 | 5 | 0.3 | 10 | 0.3 | 53 |
| I prefer not to answer | 0.1 | 2 | 0.0 | 1 | 0.1 | 3 | 0.5 | 17 | 0.1 | 4 | 0.2 | 6 | 0.1 | 33 |
| *Birth country* | | | | | | | | | | | | | | |
| Country of study | 92.6 | 3,603 | 90.4 | 3,334 | 94.5 | 3,429 | 96.3 | 3,400 | 95.6 | 4,013 | 89.4 | 2,951 | 93.3 | 20,730 |
| Other | 7.4 | 287 | 9.6 | 352 | 5.5 | 198 | 3.7 | 129 | 4.4 | 183 | 10.6 | 349 | 6.7 | 1,498 |
| *Mother's birth country* | | | | | | | | | | | | | | |
| Country of study | 87.5 | 3,405 | 88.2 | 3,250 | 86.2 | 3,126 | 95.1 | 3,355 | 94.4 | 3,959 | 81.6 | 2,693 | 89.0 | 2,440 |
| Other | 12.5 | 485 | 11.8 | 436 | 13.8 | 501 | 4.9 | 174 | 5.6 | 237 | 18.4 | 607 | 11.0 | 19,788 |
| *Father's birth country* | | | | | | | | | | | | | | |
| Country of study | 85.3 | 3,317 | 88.5 | 3,262 | 83.3 | 3,020 | 94.5 | 3,334 | 95.4 | 4,004 | 81.1 | 2,677 | 88.2 | 19,614 |
| Other | 14.7 | 573 | 11.5 | 424 | 16.7 | 607 | 5.5 | 195 | 4.6 | 192 | 18.9 | 623 | 11.8 | 2,614 |
| *Marital status* | | | | | | | | | | | | | | |
| Married or in civil union | 54.2 | 2,110 | 51.3 | 1,891 | 49.3 | 1,789 | 53.7 | 1,894 | 50.6 | 2,124 | 50.7 | 1,672 | 51.6 | 11,480 |
| Separated | 2.9 | 112 | 3.5 | 128 | 4.6 | 167 | 5.9 | 208 | 3.3 | 137 | 2.4 | 79 | 3.7 | 831 |
| Divorced or civil union dissolved | 11.0 | 429 | 6.5 | 239 | 8.5 | 309 | 9.2 | 326 | 4.9 | 206 | 6.6 | 218 | 7.8 | 1,727 |
| Widowed | 4.2 | 162 | 1.7 | 62 | 2.9 | 106 | 3.4 | 120 | 2.5 | 105 | 3.3 | 110 | 3.0 | 665 |
| Never married or in civil union | 26.9 | 1,047 | 36.1 | 1,331 | 33.1 | 1,198 | 24.8 | 874 | 36.9 | 1,548 | 34.9 | 1,153 | 32.2 | 7,151 |
| I prefer not to answer | 0.8 | 30 | 0.9 | 35 | 1.6 | 58 | 3.0 | 107 | 1.8 | 76 | 2.1 | 68 | 1.7 | 374 |
| *Minority* | | | | | | | | | | | | | | |

| | | | | | | | | | | | | | | |
|---|---|---|---|---|---|---|---|---|---|---|---|---|---|---|
| | At least 1 | 21.4 | 832 | 21.6 | 795 | 23.8 | 863 | 12.1 | 425 | 16.2 | 679 | 31.8 | 1,048 | 20.9 | 4,642 |
| | None | 77.5 | 3,015 | 77.1 | 2,842 | 73.7 | 2,672 | 85.3 | 3,011 | 83.3 | 3,496 | 66.8 | 2,206 | 77.6 | 17,242 |
| | I prefer not to answer | 1.1 | 43 | 1.3 | 49 | 2.5 | 92 | 2.6 | 93 | 0.5 | 21 | 1.4 | 46 | 1.5 | 344 |
| *Politics* | | | | | | | | | | | | | | | |
| | EPP | 20.3 | 789 | 21.1 | 778 | 8.7 | 314 | 0.0 | 0 | 7.4 | 311 | 0.0 | 0 | 9.9 | 2,192 |
| | GREENSEFA | 11.3 | 438 | 9.3 | 341 | 4.9 | 178 | 1.3 | 45 | 3.8 | 160 | 8.6 | 282 | 6.5 | 1,444 |
| | NI | 15.7 | 609 | 2.1 | 76 | 0.0 | 0 | 2.7 | 94 | 0.6 | 19 | 8.2 | 271 | 4.8 | 1,069 |
| | RenewEurope | 5.7 | 220 | 1.4 | 52 | 10.2 | 369 | 3.5 | 124 | 4.8 | 201 | 7.0 | 230 | 5.4 | 1,196 |
| | S&D | 15.2 | 597 | 27.8 | 1,025 | 6.6 | 241 | 6.2 | 217 | 14.1 | 593 | 35.0 | 1,156 | 17.2 | 3,829 |
| | Other | 6.6 | 256 | 4.5 | 167 | 4.2 | 155 | 17.9 | 633 | 2.6 | 110 | 1.6 | 54 | 6.2 | 1,375 |
| | None | 21.7 | 844 | 16.6 | 612 | 60.5 | 2,194 | 59.3 | 2,095 | 62.5 | 2,624 | 37.3 | 1, 230 | 43.2 | 9,599 |
| | I prefer not to answer | 3.5 | 137 | 17.2 | 635 | 4.9 | 176 | 9.1 | 321 | 4.2 | 178 | 2.3 | 77 | 6.8 | 1,524 |
| *Religion* | | | | | | | | | | | | | | | |
| | Atheist Agnostic | 40.7 | 1,584 | 34.9 | 1,288 | 38.7 | 1,403 | 29.7 | 1,048 | 25.8 | 1,084 | 37.9 | 1,252 | 34.5 | 7,659 |
| | Christian Catholic | 18.4 | 717 | 54.0 | 1,990 | 39.6 | 1,437 | 37.1 | 1,308 | 65.1 | 2,733 | 10.6 | 351 | 38.4 | 8,536 |
| | Christian Orthodox | 1.0 | 38 | 1.3 | 47 | 0.5 | 19 | 0.7 | 24 | 1.0 | 42 | 0.6 | 21 | 0.9 | 191 |
| | Christian Protestant | 17.4 | 676 | 0.8 | 30 | 2.0 | 74 | 15.8 | 556 | 0.5 | 21 | 29.6 | 977 | 10.5 | 2,334 |
| | Eastern Religions | 0.6 | 24 | 1.1 | 42 | 0.4 | 14 | 0.0 | 0 | 1.0 | 41 | 2.3 | 75 | 0.9 | 196 |
| | Jewish | 0.4 | 14 | 0.1 | 4 | 0.7 | 27 | 0.5 | 19 | 0.2 | 7 | 0.7 | 24 | 0.4 | 95 |
| | Muslim | 3.0 | 116 | 1.4 | 51 | 6.7 | 242 | 0.5 | 18 | 0.5 | 20 | 5.2 | 171 | 2.8 | 618 |
| | Other Christian | 14.5 | 563 | 2.7 | 101 | 4.7 | 169 | 3.5 | 122 | 2.6 | 109 | 8.0 | 263 | 6.0 | 1,327 |
| | Other non-Christian | 0.6 | 25 | 1.1 | 39 | 2.2 | 79 | 2.9 | 102 | 0.9 | 37 | 1.5 | 49 | 1.5 | 331 |
| | I prefer not to answer | 3.4 | 133 | 2.6 | 94 | 4.5 | 163 | 9.4 | 332 | 2.4 | 102 | 3.5 | 117 | 4.2 | 941 |
| *Occupation* | | | | | | | | | | | | | | | |
| | Employed | 53.1 | 2,066 | 50.3 | 1,853 | 51.4 | 1,866 | 60.7 | 2,141 | 41.7 | 1,748 | 53.1 | 1,754 | 51.4 | 11,428 |
| | Self-employed | 6.4 | 248 | 7.0 | 259 | 6.1 | 222 | 6.6 | 233 | 12.6 | 527 | 7.2 | 237 | 7.8 | 1,726 |
| | Employed with on-call, seasonal, occasional work | 1.9 | 76 | 9.0 | 331 | 3.3 | 119 | 3.7 | 131 | 3.7 | 154 | 3.4 | 111 | 4.1 | 922 |
| | Student | 3.9 | 152 | 8.6 | 317 | 5.8 | 211 | 4.8 | 170 | 6.3 | 267 | 3.8 | 127 | 5.7 | 1,244 |

| | | | | | | | | | | | | | | |
|---|---|---|---|---|---|---|---|---|---|---|---|---|---|---|
| | Unemployed | 5.5 | 213 | 11.4 | 421 | 7.1 | 257 | 6.1 | 215 | 11.4 | 477 | 8.3 | 273 | 8.3 | 1,856 |
| | Inactive (e.g., retired, homemaker) | 25.5 | 993 | 11.8 | 436 | 22.9 | 829 | 14.9 | 525 | 21.6 | 909 | 20.8 | 686 | 19.7 | 4,378 |
| | Other | 3.2 | 124 | 1.5 | 56 | 2.9 | 104 | 2.5 | 87 | 2.4 | 100 | 2.9 | 95 | 2.5 | 566 |
| | I prefer not to answer | 0.5 | 18 | 0.4 | 13 | 0.5 | 19 | 0.7 | 27 | 0.3 | 14 | 0.5 | 17 | 0.5 | 108 |
| *Cohabitant* | | | | | | | | | | | | | | | |
| | 0 | 31.1 | 1,210 | 12.6 | 465 | 23.0 | 833 | 17.3 | 610 | 16.4 | 689 | 28.4 | 935 | 21.3 | 4,742 |
| | 1 | 24.2 | 940 | 21.4 | 790 | 29.4 | 1,068 | 18.3 | 646 | 29.1 | 1,223 | 30.3 | 1,000 | 25.5 | 5,667 |
| | 2 | 23.6 | 917 | 27.8 | 1,025 | 21.5 | 779 | 25.6 | 902 | 26.8 | 1,123 | 17.4 | 575 | 23.9 | 5,321 |
| | 3-5 | 20.3 | 791 | 37.3 | 1,374 | 25.3 | 919 | 36.7 | 1,296 | 27.2 | 1,141 | 23.2 | 766 | 28.3 | 6,287 |
| | >5 | 0.8 | 32 | 0.9 | 32 | 0.8 | 28 | 2.1 | 75 | 0.5 | 20 | 0.7 | 24 | 1.0 | 211 |
| *Non-cohabitant family* | | | | | | | | | | | | | | | |
| | 0 | 25.3 | 983 | 26.8 | 988 | 35.6 | 1,292 | 19.4 | 685 | 43.1 | 1,807 | 44.1 | 1,456 | 32.4 | 7,211 |
| | 1 | 11.0 | 430 | 12.4 | 457 | 11.9 | 431 | 14.6 | 514 | 14.4 | 605 | 12.9 | 424 | 12.9 | 2,861 |
| | 2 | 17.0 | 661 | 16.8 | 619 | 14.1 | 510 | 20.7 | 732 | 15.0 | 630 | 12.9 | 427 | 16.1 | 3,579 |
| | 3-5 | 35.5 | 1,381 | 34.3 | 1,266 | 28.6 | 1,036 | 36.1 | 1,275 | 22.9 | 960 | 23.0 | 758 | 30.0 | 6,676 |
| | >5 | 11.2 | 435 | 9.7 | 356 | 9.8 | 358 | 9.2 | 323 | 4.6 | 194 | 7.1 | 235 | 8.6 | 1,901 |
| **Economic** | | | | | | | | | | | | | | | |
| *Income* | | | | | | | | | | | | | | | |
| | Quintile 1 | 18.6 | 722 | 14.0 | 514 | 16.3 | 590 | 14.0 | 496 | 14.6 | 612 | 15.1 | 497 | 15.4 | 3,431 |
| | Quintile 2 | 19.9 | 776 | 25.3 | 932 | 17.0 | 617 | 13.9 | 490 | 22.9 | 961 | 22.4 | 739 | 20.3 | 4,515 |
| | Quintile 3 | 22.9 | 891 | 22.0 | 813 | 16.4 | 593 | 12.7 | 449 | 22.7 | 954 | 23.2 | 764 | 20.1 | 4,464 |
| | Quintile 4 | 19.9 | 773 | 23.2 | 855 | 18.3 | 664 | 15.9 | 560 | 18.8 | 788 | 18.9 | 624 | 19.2 | 4,264 |
| | Quintile 5 | 13.9 | 541 | 11.8 | 437 | 28.5 | 1,035 | 37.5 | 1,323 | 13.4 | 562 | 16.0 | 529 | 19.9 | 4,427 |
| | I prefer not to answer | 4.8 | 187 | 3.7 | 135 | 3.5 | 128 | 6.0 | 211 | 7.6 | 319 | 4.4 | 147 | 5.1 | 1,127 |
| **Health** | | | | | | | | | | | | | | | |
| *Perceived health status* | | | | | | | | | | | | | | | |
| | Very good | 12.2 | 473 | 15.1 | 558 | 15.7 | 568 | 10.9 | 384 | 9.6 | 403 | 17.2 | 566 | 13.3 | 2,952 |
| | Good | 45.5 | 1,769 | 48.4 | 1,785 | 58.5 | 2,121 | 46.9 | 1,656 | 47.8 | 2,003 | 45.4 | 1,498 | 48.7 | 10,832 |
| | Fair | 32.3 | 1,257 | 29.1 | 1,072 | 19.8 | 719 | 34.7 | 1,223 | 36.1 | 1,516 | 28.9 | 952 | 30.3 | 6,739 |
| | Bad | 8.3 | 322 | 6.3 | 231 | 4.8 | 175 | 6.2 | 220 | 5.2 | 216 | 6.5 | 216 | 6.2 | 1,380 |

| | | | | | | | | | | | | | | |
|---|---|---|---|---|---|---|---|---|---|---|---|---|---|---|
| | Very bad | 1.3 | 52 | 0.9 | 33 | 0.9 | 33 | 1.0 | 36 | 1.1 | 48 | 1.8 | 60 | 1.2 | 262 |
| | I prefer not to answer | 0.4 | 17 | 0.2 | 7 | 0.3 | 11 | 0.3 | 10 | 0.2 | 10 | 0.2 | 8 | 0.3 | 63 |
| *Medical conditions* | | | | | | | | | | | | | | | |
| | At least 1 | 47.2 | 1,834 | 34.6 | 1,277 | 34.2 | 1,241 | 38.9 | 1,372 | 33.4 | 1,403 | 33.1 | 1,092 | 37.0 | 8,219 |
| | None | 51.1 | 1,989 | 64.0 | 2,358 | 63.8 | 2,314 | 59.1 | 2,086 | 65.2 | 2,734 | 64.9 | 2,143 | 61.3 | 13,624 |
| | I prefer not to answer | 1.7 | 67 | 1.4 | 51 | 2.0 | 72 | 2.0 | 71 | 1.4 | 59 | 2.0 | 65 | 1.7 | 385 |
| *Cohabitants' medical conditions* | | | | | | | | | | | | | | | |
| | Yes | 21.8 | 849 | 27.6 | 1,017 | 16.4 | 597 | 30.3 | 1,069 | 22.6 | 948 | 17.4 | 575 | 22.8 | 5,055 |
| | No | 43.9 | 1,707 | 56.4 | 2,080 | 58.2 | 2,110 | 46.6 | 1,646 | 58.0 | 2,434 | 51.2 | 1,690 | 52.6 | 11,667 |
| | I don't know | 2.1 | 83 | 2.9 | 107 | 1.8 | 66 | 5.0 | 177 | 2.1 | 86 | 2.1 | 67 | 2.6 | 586 |
| | None | 31.1 | 1,210 | 12.6 | 465 | 23.0 | 833 | 17.3 | 610 | 16.4 | 689 | 28.3 | 935 | 21.3 | 4,742 |
| | I prefer not to answer | 1.1 | 41 | 0.5 | 17 | 0.6 | 21 | 0.8 | 27 | 0.9 | 39 | 1.0 | 33 | 0.7 | 178 |

**Table S4** Filtering and flagging criteria Screening filters and quality flags were applied to the raw survey dataset during or after the data collection to minimise fraudulent and unreliable responses and ensure high data quality.

| **Filtering criteria** | **Details** | **#** |
|---|---|---|
| Automated and duplicated responses | • Responses classified as bots by the data collection platform Qualtrics<br>• Among responses with at least one duplicate field among respondent ID, recontact email, and social media user handles, or with identical answers across all non-drop-down questions presented to all respondents:<br> ○ if all responses had overlapping survey completion timestamps, all were dropped;<br> ○ if responses had non-overlapping timestamps, earliest response was retained, later response(s) were dropped | n=5,172 |
| Unrealistic survey completion time | • Responses with survey completion time below a set threshold. The threshold was defined as one-third of the median completion time for their reference group. Reference groups were defined based on shared characteristics, including country, number of COVID-19 infections, number of COVID-19 vaccine doses, household size, and number of non-cohabiting contacts | n=92 |
| Patterned or non-differentiated responses | • Responses exhibiting straight lining, *i.e.*, selecting the same answer option for all socio-demographic questions or across all items in tables or carousel-style questions | n=175 |
| Implausible relationship-age pairings | • Responses indicating unrealistic relationship-age pairings, *i.e.* not meeting the following criteria:<br> ○ Parents: min. age = age of the respondent + 14 years<br> ○ Grandparents: min. age = age of the respondent + 28 years<br> ○ Children: min. age = age of the respondent – 14 years<br> ○ Partners: min. age = 14 years old | n=2,746 |
| Inconsistent open text | • Open text responses inconsistent with previous answers | n=70 |
| Invalid geographic residence | • Responses indicating residence in extraterritorial domains | n=4 |
| **Flagging criteria** | **Details** | **#** |
| Implausible COVID-19 infection or vaccination dates | • Wrong temporal order of reported multiple infections or vaccinations<br>• Dates following survey time<br>• Vaccinations received before December 2020 | n=1,141 |
| Implausible social contact numbers | • Respondents reporting a number of discussion or in-person contacts in one of their social groups that is higher than the maximum number of people reported earlier in that specific social group | n=10,118 |

| | | |
|---|---|---|
| Implausible school attendance | • Respondents compiling the questionnaire on a Monday and stating in-presence attendance at school on the day prior to the survey | n=160 |
| Implausible values | • Respondents reporting decimal or negative values or matching specific regular expressions when asked to indicate the number of people in a specific group | n=61 |
| Single-dose vaccine regimen | • Respondents declaring in open-text questions that they received the Johnson&Johnson vaccine, which follows a single-dose schedule differing from the multi-dose regimens of other vaccines included in the survey | n=50 |

**Table S5** COVID-19 vaccine uptake by level of willingness to vaccinate Proportion of respondents taking up 0, 1, 2, 3, or 4+ doses of COVID-19 vaccine, by country and stated willingness to get vaccinated when the first dose was offered to them. The table excludes respondents who i) indicated that COVID-19 vaccination was mandatory for them (DE n=573; ES n=540; FR n=704; HU n=517; IT n=637; UK n=442); ii) indicated either wanting the mandatory vaccination certificate (DE n=904; ES n=783; FR n=1,436; HU n=638; IT n=1,305; UK n=560); iii) provided free-text responses as main reasons for vaccination (DE n=164; ES n=59; FR n=42; HU n=107; IT n=102; UK n=48); iv) reported a medical condition that prevented them from getting vaccinated (DE n=15; ES n=4; FR n=12; HU n=24; IT n =15; UK n =2); v) stopped vaccinating because they experienced COVID-19 illness (DE n=526; ES n=243; FR n=479; HU n=140; IT n=646; UK n=203). Respondents who were administered 1 (n=21), 2 (n=21), or 3 (n=5) Johnson & Johnson vaccine doses were recoded as having received a total of 2, 3 or 4 doses, respectively.

| # of vaccine doses taken | Respondents' stated willingness to get vaccinated against COVID-19 when the first dose was offered to them (%) | | | | | All |
|---|---|---|---|---|---|---|
| | Not at all | Not very | Quite | Very | Extremely | |
| ***DE*** | | | | | | |
| 0 | 89.0 | 32.9 | 5.9 | 1.9 | 1.5 | 26.6 |
| 1 | 1.2 | 6.7 | 4.7 | 2.4 | 0.6 | 2.5 |
| 2 | 4.9 | 27.5 | 34.5 | 20.7 | 8.1 | 16.6 |
| 3 | 4.4 | 29.3 | 44.7 | 46.2 | 39.0 | 32.4 |
| 4+ | 0.5 | 3.6 | 10.2 | 28.8 | 50.8 | 21.9 |
| ***ES*** | | | | | | |
| 0 | 78.2 | 19.3 | 3.2 | 1.5 | 0.7 | 14.0 |
| 1 | 3.9 | 7.6 | 3.2 | 1.8 | 0.5 | 2.9 |
| 2 | 10.1 | 36.7 | 40.7 | 26.8 | 17.8 | 27.5 |
| 3 | 6.2 | 32.0 | 40.1 | 49.3 | 41.9 | 37.4 |
| 4+ | 1.6 | 4.4 | 12.8 | 20.6 | 39.1 | 18.2 |

| | | | | | | |
|---|---|---|---|---|---|---|
| ***FR*** | | | | | | |
| 0 | 86.1 | 14.4 | 9.1 | 4.5 | 1.7 | 35.4 |
| 1 | 1.8 | 7.5 | 3.3 | 4.8 | 0.4 | 3.3 |
| 2 | 4.9 | 34.7 | 28.0 | 16.8 | 7.3 | 15.8 |
| 3 | 5.8 | 26.9 | 38.2 | 35.0 | 34.2 | 23.9 |
| 4+ | 1.4 | 16.5 | 21.4 | 38.9 | 56.4 | 21.6 |
| ***HU*** | | | | | | |
| 0 | 88.6 | 28.0 | 13.1 | 4.8 | 2.7 | 44.9 |
| 1 | 1.5 | 8.0 | 4.5 | 1.1 | 0.6 | 2.6 |
| 2 | 4.6 | 33.2 | 35.4 | 20.3 | 15.2 | 17.2 |
| 3 | 5.1 | 29.9 | 43.5 | 66.3 | 58.3 | 30.7 |
| 4+ | 0.2 | 0.9 | 3.5 | 7.5 | 23.2 | 4.6 |
| ***IT*** | | | | | | |
| 0 | 88.2 | 23.7 | 6.0 | 0.5 | 0.4 | 20.4 |
| 1 | 0.6 | 5.2 | 1.4 | 0.6 | 0.0 | 1.1 |
| 2 | 3.9 | 25.2 | 22.9 | 13.9 | 8.6 | 13.9 |
| 3 | 6.7 | 42.1 | 59.7 | 58.0 | 51.7 | 45.7 |
| 4+ | 0.6 | 3.8 | 10.0 | 27.0 | 39.3 | 18.9 |
| ***UK*** | | | | | | |
| 0 | 89.3 | 37.5 | 10.4 | 4.5 | 1.2 | 20.0 |

| | | | | | | |
|---|---|---|---|---|---|---|
| 1 | 5.0 | 12.0 | 10.6 | 5.1 | 1.5 | 5.3 |
| 2 | 2.5 | 32.0 | 36.1 | 27.1 | 11.6 | 19.8 |
| 3 | 2.3 | 15.0 | 32.4 | 37.2 | 35.8 | 28.6 |
| 4+ | 0.9 | 3.5 | 10.5 | 26.1 | 49.9 | 26.3 |
| ***All countries*** | | | | | | |
| 0 | 87.2 | 25.8 | 7.5 | 2.7 | 1.2 | 26.7 |
| 1 | 2.0 | 7.9 | 4.4 | 2.6 | 0.7 | 2.9 |
| 2 | 4.9 | 31.9 | 33.5 | 21.4 | 11.5 | 18.7 |
| 3 | 5.1 | 29.1 | 43.7 | 48.7 | 42.3 | 33.4 |
| 4+ | 0.8 | 5.3 | 10.9 | 24.6 | 44.3 | 18.3 |

**Table S6** COVID-19 vaccination timeliness Average number of months between the time point (MM/YYY) when the COVID-19 vaccine was first offered to each specific age group and when vaccinated individuals in that group received their first dose by country. "High" willingness includes respondents who indicated they were very or extremely willing to get vaccinated at the time the first dose was offered to them; "medium" willingness includes individuals who indicated they were quite willing to get vaccinated at the same time point; "low" willingness includes those who were not very or not at all willing to get vaccinated. The table excludes respondents who i) indicated that COVID-19 vaccination was mandatory for them (DE n=573; ES n=540; FR n=704; HU n=517; IT n=637; UK n=442); ii) indicated wanting the mandatory vaccination certificate (DE n=904; ES n=783; FR n=1,436; HU n=638; IT n=1,305; UK n=560); iii) provided free-text responses as main reasons for vaccination (DE n=164; ES n=59; FR n=42; HU n=107; IT n=102; UK n=48); indicated uptake dates preceding the vaccine offer to their age category (DE n=574; ES n=835; FR n=447; HU n=430; IT n=901; UK n=683); were administered the Johnson&Johnson vaccine, which was offered to the population with a delay (DE n=11; ES n=8; FR n=6; HU n=17; IT n=6; UK n=2).

| **Willingness** | **Age group** | | | | | **All** |
|---|---|---|---|---|---|---|
| | **18-29 years** | **30-39 years** | **40-49 years** | **50-59 years** | **60+ years** | |
| ***DE*** | | | | | | |
| High | 5.0 | 3.8 | 3.2 | 2.4 | 3.2 | 3.4 |
| Medium | 6.2 | 5.4 | 4.1 | 5.3 | 3.5 | 5.0 |
| Low | 7.5 | 5.2 | 6.1 | 5.4 | 5.6 | 6.1 |
| All | 5.8 | 4.5 | 3.9 | 3.3 | 3.5 | 4.0 |
| ***ES*** | | | | | | |
| High | 5.2 | 4.2 | 2.7 | 2.5 | 2.8 | 3.3 |
| Medium | 4.7 | 3.3 | 4.0 | 3.4 | 3.8 | 3.8 |
| Low | 7.5 | 4.9 | 3.7 | 2.0 | 3.6 | 4.6 |
| All | 5.5 | 4.1 | 3.2 | 2.7 | 3.2 | 3.7 |
| ***FR*** | | | | | | |
| High | 6.9 | 5.2 | 4.7 | 4.9 | 4.4 | 5.0 |

| | | | | | | |
|---|---|---|---|---|---|---|
| Medium | 4.9 | 7.2 | 5.8 | 3.6 | 4.7 | 5.3 |
| Low | 6.0 | 7.3 | 7.2 | 5.0 | 5.6 | 6.2 |
| All | 5.9 | 6.4 | 5.7 | 4.6 | 4.7 | 5.4 |
| ***HU*** | | | | | | |
| High | 5.2 | 1.3 | 1.7 | 2.2 | 3.1 | 2.5 |
| Medium | 6.8 | 3.4 | 2.6 | 4.7 | 4.0 | 4.2 |
| Low | 7.6 | 4.8 | 4.1 | 4.9 | 3.6 | 5.3 |
| All | 6.6 | 2.9 | 2.5 | 3.4 | 3.4 | 4.0 |
| ***IT*** | | | | | | |
| High | 1.7 | 2.3 | 1.8 | 1.5 | 2.4 | 2.0 |
| Medium | 4.9 | 3.6 | 3.7 | 3.6 | 4.0 | 3.9 |
| Low | 6.6 | 5.3 | 3.7 | 3.5 | 3.6 | 4.1 |
| All | 2.8 | 3.1 | 2.6 | 2.4 | 2.9 | 3.0 |
| ***UK*** | | | | | | |
| High | 5.7 | 6.3 | 3.1 | 3.3 | 2.3 | 3.9 |
| Medium | 7.7 | 7.1 | 4.5 | 3.5 | 3.8 | 5.7 |
| Low | 6.4 | 6.5 | 5.5 | 6.2 | 2.9 | 5.7 |
| All | 6.4 | 6.5 | 3.8 | 3.6 | 2.5 | 5.0 |
| ***All countries*** | | | | | | |
| High | 4.7 | 3.9 | 2.7 | 2.7 | 2.7 | 3.0 |
| Medium | 6.0 | 4.6 | 3.8 | 3.9 | 4.0 | 4.0. |

| | | | | | | |
|---|---|---|---|---|---|---|
| Low | 7.1 | 5.6 | 4.9 | 4.4 | 4.3 | 5.0 |
| All | 5.6 | 3.9 | 3.4 | 3.2 | 3.3 | 4.0 |

**Table S7** Radar plot variables Survey questions involving Likert answer scales, corresponding to the variables plotted in **Fig. 3**. The variables are stratified by *Capability*, *Opportunity* and *Motivation* behaviour (COM-B) model dimensions. In the *Opportunity* section, responses only included those from respondents who previously indicated the presence of each given social group.

| COM-B dimension | Variable | Survey question | Answer scale |
|---|---|---|---|
| Capability | Booking ability | To what extent do you feel able to independently book vaccination appointments for yourself or your family members? | 5-point ability scale |
| | Access to vaccination | How easy is it for you to independently reach the nearest vaccination facility? | 5-point ability scale |
| Opportunity | [Social group]'s opinion | To what extent do people from the following groups agree or disagree with the statements below, approximately?<br>*Vaccines are decisive for the protection of human health* | 7-point agreement scale |
| Motivation | Risk of vaccine | In your opinion, how likely are you to experience health consequences (such as severe side effects) if you receive the following vaccines? | 5-point likelihood scale |
| | Fear of disease | How much do you fear each of the following illnesses? | 5-point emotional scale |
| | Fear of vaccine | How much do you fear each of the following vaccines? | 5-point emotional scale |
| | Confidence | Please evaluate how much you disagree or agree with the following statements:<br>*I am completely confident that vaccines are safe* | 7-point agreement scale |
| | Complacency | Please evaluate how much you disagree or agree with the following statements:<br>*Vaccination is unnecessary because vaccine-preventable diseases are not common anymore* | 7-point agreement scale |
| | Calculation | Please evaluate how much you disagree or agree with the following statements:<br>*When I think about getting vaccinated, I weigh benefits and risks to make the best decision possible* | 7-point agreement scale |
| | Collective responsibility | Please evaluate how much you disagree or agree with the following statements:<br>*When everyone is vaccinated, I don't have to get vaccinated* | 7-point agreement scale |
| | Trust in [institution] | How much do you trust the information about vaccines/vaccination programs shared by the following people or institutions? | 5-point trust scale |

**Table S8** Radar plot values Weighted means of Likert scale values chosen by participants in response to selected survey questions across *Capability*, *Opportunity* and *Motivation* behaviour (COM-B) model dimensions, stratified by country and respondents' COVID-19 or influenza vaccination status. Values for the COVID-19 columns exclude respondents who i) indicated that COVID-19 vaccination was mandatory for them (DE n=573; ES n=540; FR n=704; HU n=517; IT n=637; UK n=442); ii) indicated wanting the mandatory vaccination certificate (DE n=904; ES n=783; FR n=1,436; HU n=638; IT n=1,305; UK n=560); iii) provided free-text responses as main reasons for vaccination (DE n=164; ES n=59; FR n=42; HU n=107; IT n=102; UK n=48); iv) reported a medical condition that prevented them from getting vaccinated (DE n=15; ES n=4; FR n=12; HU n=24; IT n =15; UK n =2). Weighted means for partners', siblings', parents', and friends' opinions only include those from participants who previously reported the presence of each given social group. For the calculations, 7-point scales were transformed into 5-point scales by pooling the three central options ("Slightly disagree", "Neutral", and "Slightly agree").

| | COVID-19 | | Influenza | |
|---|---|---|---|---|
| **Variable** | **Vaccinated*** | **Unvaccinated** | **Vaccinated**** | **Unvaccinated** |
| ***DE*** | | | | |
| *Capability* | | | | |
| Booking ability | 4.4 | 3.9 | 4.5 | 4.2 |
| Access to vaccination | 4.1 | 3.6 | 4.2 | 3.9 |
| *Opportunity* | | | | |
| Partners' opinion | 3.4 | 2.6 | 3.5 | 3.0 |
| Siblings' opinion | 3.2 | 2.7 | 3.2 | 3.0 |
| Parents' opinion | 3.2 | 2.6 | 3.3 | 3.0 |
| Friends' opinion | 3.0 | 2.6 | 3.1 | 2.9 |
| Coworkers' opinion | 3.0 | 2.8 | 3.0 | 2.9 |
| *Motivation* | | | | |
| Risk of vaccine | 2.2 | 3.5 | 2.3 | 2.7 |
| Fear of vaccine | 1.9 | 3.7 | 2.0 | 2.6 |
| Fear of disease | 2.4 | 1.6 | 2.4 | 1.9 |
| Confidence | 3.4 | 2.1 | 3.4 | 2.8 |
| Complacency | 1.9 | 2.8 | 1.8 | 2.3 |
| Calculation | 3.4 | 3.7 | 3.4 | 3.5 |
| Collective responsibility | 2.0 | 2.9 | 1.9 | 2.4 |
| Trust in religious leader | 1.4 | 1.3 | 1.4 | 1.3 |
| Trust in national government | 2.6 | 1.6 | 2.7 | 2.1 |
| Trust in family doctor | 3.7 | 2.4 | 3.8 | 3.1 |

| | | | | |
|---|---|---|---|---|
| Trust in national health authorities | 3.1 | 1.8 | 3.1 | 2.6 |
| Trust in WHO | 3.1 | 1.8 | 3.1 | 2.6 |
| ***ES*** | | | | |
| *Capability* | | | | |
| Booking ability | 3.9 | 2.9 | 4.0 | 3.6 |
| Access to vaccination | 3.9 | 3.5 | 4.0 | 3.9 |
| *Opportunity* | | | | |
| Partners' opinion | 3.9 | 2.8 | 4.0 | 3.6 |
| Siblings' opinion | 3.7 | 2.8 | 3.7 | 3.5 |
| Parents' opinion | 3.8 | 3.0 | 3.8 | 3.7 |
| Friends' opinion | 3.5 | 2.9 | 3.4 | 3.3 |
| Coworkers' opinion | 3.3 | 3.0 | 3.3 | 3.3 |
| *Motivation* | | | | |
| Risk of vaccine | 2.2 | 3.1 | 2.2 | 2.4 |
| Fear of vaccine | 1.9 | 3.2 | 1.9 | 2.3 |
| Fear of disease | 2.4 | 1.8 | 2.5 | 2.1 |
| Vaccine confidence | 3.8 | 2.4 | 3.9 | 3.4 |
| Complacency | 1.9 | 2.8 | 1.9 | 2.2 |
| Calculation | 3.5 | 3.8 | 3.6 | 3.6 |
| Collective responsibility | 1.9 | 2.8 | 1.9 | 2.2 |
| Trust in religious leader | 1.6 | 1.5 | 1.6 | 1.5 |
| Trust in national government | 2.6 | 1.9 | 2.7 | 2.3 |
| Trust in family doctor | 3.7 | 2.5 | 3.8 | 3.3 |
| Trust in national health authorities | 3.3 | 2.3 | 3.4 | 3.0 |
| Trust in WHO | 3.3 | 2.1 | 3.4 | 3.6 |
| ***FR*** | | | | |
| *Capability* | | | | |
| Booking ability | 4.1 | 3.4 | 4.1 | 3.9 |
| Access to vaccination | 4.0 | 3.4 | 4.1 | 3.8 |
| *Opportunity* | | | | |

| | | | | |
|---|---|---|---|---|
| Partners' opinion | 3.6 | 2.6 | 3.5 | 3.2 |
| Siblings' opinion | 3.3 | 2.7 | 3.3 | 3.1 |
| Parents' opinion | 3.3 | 2.8 | 3.2 | 3.1 |
| Friends' opinion | 3.1 | 2.7 | 3.1 | 3.0 |
| Coworkers' opinion | 3.1 | 2.7 | 3.0 | 2.9 |
| *Motivation* | | | | |
| Risk of vaccine | 2.1 | 3.3 | 2.3 | 2.6 |
| Fear of vaccine | 2.0 | 3.6 | 2.1 | 2.7 |
| Fear of disease | 2.4 | 1.7 | 2.4 | 1.9 |
| Vaccine confidence | 3.3 | 2.2 | 3.4 | 2.8 |
| Complacency | 2.1 | 2.9 | 2.1 | 2.5 |
| Calculation | 3.2 | 3.5 | 3.2 | 3.4 |
| Collective responsibility | 2.1 | 2.8 | 2.1 | 2.4 |
| Trust in religious leader | 1.7 | 1.6 | 1.7 | 1.5 |
| Trust in national government | 2.68 | 1.7 | 2.8 | 2.2 |
| Trust in family doctor | 3.7 | 2.6 | 3.8 | 3.2 |
| Trust in national health authorities | 3.3 | 2.1 | 3.3 | 2.7 |
| Trust in WHO | 3.3 | 2.1 | 3.3 | 2.7 |
| ***HU*** | | | | |
| *Capability* | | | | |
| Booking ability | 4.4 | 3.7 | 4.4 | 4.1 |
| Access to vaccination | 4.1 | 3.5 | 4.0 | 3.9 |
| *Opportunity* | | | | |
| Partners' opinion | 3.3 | 2.6 | 3.3 | 2.8 |
| Siblings' opinion | 3.0 | 2.6 | 3.1 | 2.8 |
| Parents' opinion | 3.1 | 2.6 | 3.0 | 2.8 |
| Friends' opinion | 2.9 | 2.6 | 2.9 | 2.7 |
| Coworkers' opinion | 2.9 | 2.6 | 2.9 | 2.7 |
| *Motivation* | | | | |
| Risk of vaccine | 2.7 | 3.8 | 2.9 | 3.3 |

| | | | | |
|---|---|---|---|---|
| Fear of vaccine | 1.8 | 3.4 | 2.1 | 2.6 |
| Fear of disease | 2.1 | 1.6 | 2.1 | 1.8 |
| Vaccine confidence | 3.1 | 2.0 | 3.0 | 2.5 |
| Complacency | 2.1 | 2.9 | 2.0 | 2.5 |
| Calculation | 3.5 | 3.6 | 3.6 | 3.6 |
| Collective responsibility | 2.1 | 3.0 | 2.1 | 2.5 |
| Trust in religious leader | 1.7 | 1.5 | 1.7 | 1.5 |
| Trust in national government | 2.3 | 1.6 | 2.2 | 1.9 |
| Trust in family doctor | 3.5 | 2.4 | 3.4 | 2.8 |
| Trust in national health authorities | 3.0 | 2.1 | 3.0 | 2.5 |
| Trust in WHO | 3.2 | 2.1 | 3.2 | 2.6 |
| ***IT*** | | | | |
| *Capability* | | | | |
| Booking ability | 4.0 | 3.3 | 4.0 | 3.8 |
| Access to vaccination | 3.9 | 3.4 | 3.9 | 3.8 |
| *Opportunity* | | | | |
| Partners' opinion | 3.8 | 2.5 | 3.8 | 3.2 |
| Siblings' opinion | 3.5 | 2.9 | 3.5 | 3.2 |
| Parents' opinion | 3.6 | 2.8 | 3.7 | 3.3 |
| Friends' opinion | 3.1 | 2.7 | 3.1 | 3.0 |
| Coworkers' opinion | 3.0 | 2.8 | 3.1 | 2.9 |
| *Motivation* | | | | |
| Risk of vaccine | 2.1 | 3.5 | 2.2 | 2.6 |
| Fear of vaccine | 2.0 | 3.9 | 2.1 | 2.6 |
| Fear of disease | 2.4 | 1.7 | 2.4 | 2.1 |
| Vaccine confidence | 3.7 | 1.9 | 3.7 | 3.0 |
| Complacency | 2.0 | 3.0 | 2.0 | 2.4 |
| Calculation | 3.6 | 4.0 | 3.7 | 3.7 |
| Collective responsibility | 1.9 | 3.0 | 2.0 | 2.3 |
| Trust in religious leader | 1.7 | 1.3 | 1.8 | 1.5 |

| | | | | |
|---|---|---|---|---|
| Trust in national government | 2.6 | 1.5 | 2.6 | 2.2 |
| Trust in family doctor | 3.5 | 2.1 | 3.5 | 3.0 |
| Trust in national health authorities | 3.3 | 1.7 | 3.2 | 2.7 |
| Trust in WHO | 3.3 | 1.6 | 3.2 | 2.7 |
| ***UK*** | | | | |
| *Capability* | | | | |
| Booking ability | 4.0 | 3.1 | 4.0 | 3.6 |
| Access to vaccination | 3.9 | 3.2 | 4.0 | 3.6 |
| *Opportunity* | | | | |
| Partners' opinion | 3.7 | 2.8 | 3.8 | 3.1 |
| Siblings' opinion | 3.4 | 2.9 | 3.6 | 3.1 |
| Parents' opinion | 3.7 | 2.9 | 3.7 | 3.3 |
| Friends' opinion | 3.3 | 2.8 | 3.4 | 3.1 |
| Coworkers' opinion | 3.2 | 2.8 | 3.2 | 3.0 |
| *Motivation* | | | | |
| Risk of vaccine | 2.2 | 2.9 | 2.2 | 2.5 |
| Fear of vaccine | 1.8 | 3.1 | 1.8 | 2.4 |
| Fear of disease | 2.5 | 1.9 | 2.6 | 2.0 |
| Vaccine confidence | 3.6 | 2.3 | 3.7 | 3.0 |
| Complacency | 1.9 | 2.9 | 1.9 | 2.4 |
| Calculation | 3.5 | 3.5 | 3.6 | 3.5 |
| Collective responsibility | 1.9 | 3.0 | 1.9 | 2.4 |
| Trust in religious leader | 1.9 | 1.8 | 1.9 | 1.7 |
| Trust in national government | 2.8 | 1.9 | 2.9 | 2.3 |
| Trust in family doctor | 3.7 | 2.6 | 3.8 | 3.1 |
| Trust in national health authorities | 3.6 | 2.4 | 3.6 | 3.0 |
| Trust in WHO | 3.5 | 2.3 | 3.5 | 2.9 |

* Includes those who took up at least one dose of COVID-19 vaccine ever
** Includes those who took up at least one dose of influenza vaccine in the five years preceding the survey

**Table S9** Average number of daily discussion and in-person contacts by age group and country The number of weekly discussion contacts as assessed in the survey was recalculated as daily. The maximum number of both discussion and in-person contacts was capped at 30 daily contacts, with values >30 being imposed to 30. This table excludes respondents flagged as reporting implausible contact numbers (DE n=1,452; ES n=1,725; FR n=1,395; HU n=1,561; IT n=1,780; UK n=1,180).

| **Age group** | **DE** | **ES** | **FR** | **HU** | **IT** | **UK** |
|---|---|---|---|---|---|---|
| *Discussion contacts* | | | | | | |
| 18-29 | 1.4 | 2.8 | 1.7 | 2.2 | 2.1 | 2.5 |
| 30-39 | 1.6 | 2.4 | 2.4 | 2.3 | 2.9 | 2.5 |
| 40-49 | 1.7 | 2.4 | 2.0 | 2.1 | 2.9 | 2.5 |
| 50-59 | 1.6 | 2.0 | 1.7 | 1.5 | 2.2 | 1.4 |
| 60+ | 0.7 | 0.5 | 0.8 | 0.6 | 0.9 | 1.2 |
| *In-person contacts* | | | | | | |
| 18-29 | 7.8 | 12.7 | 10.9 | 10.7 | 9.9 | 11.3 |
| 30-39 | 7.3 | 10.3 | 11.1 | 11.2 | 11.3 | 9.6 |
| 40-49 | 7.3 | 10.1 | 10.7 | 9.6 | 10.8 | 9.0 |
| 50-59 | 6.4 | 8.5 | 8.1 | 6.8 | 8.9 | 5.9 |
| 60+ | 3.3 | 2.7 | 4.3 | 2.2 | 4.1 | 4.4 |

#### Supplementary Figures

**Fig. S1** Correlation between the self-reported behavioural outcomes assessed in this survey From top to bottom: i) number of COVID-19 vaccine doses received; ii) number of influenza vaccine doses received during the five years prior to the survey; iii) mask use when experiencing respiratory symptoms (at the time of the survey); iv) self-isolation when experiencing respiratory symptoms (at the time of the survey); v) mask use in crowded places when the epidemic pressure is high (at the time of the survey).

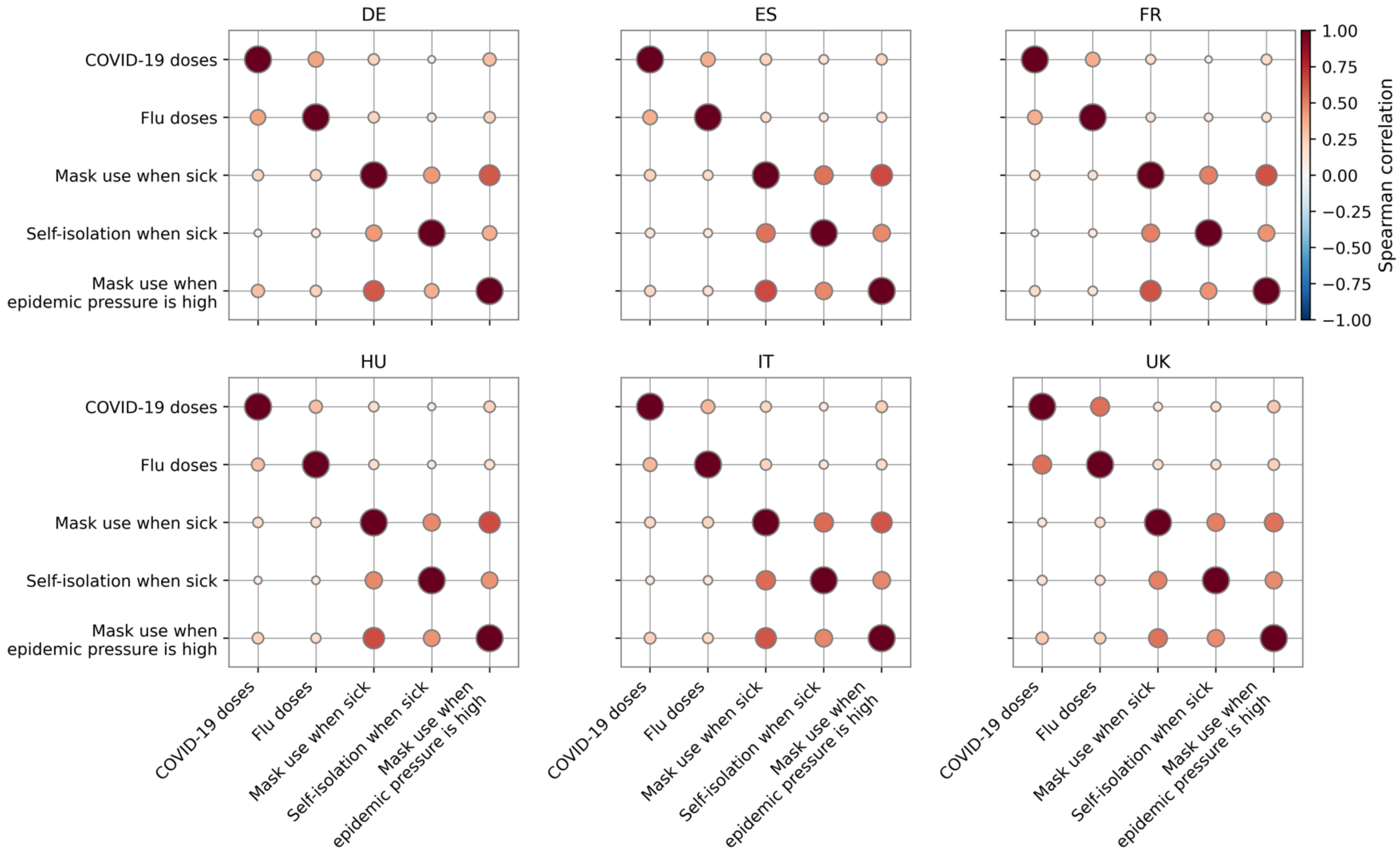

**Fig. S2** Systematic mapping of factors potentially associated with influenza vaccination behaviour Weighted means of Likert scale values chosen by participants in response to selected survey questions across *Capability*, *Opportunity* and *Motivation* behaviour (COM-B) model dimensions, stratified by country and respondents' influenza vaccination status. Vaccinated individuals include those who took up at least one dose of influenza vaccine in the five years preceding the survey. In the *Opportunity* section, responses only include those from participants who previously reported the presence of each given social group. Questionnaire items corresponding to the plotted variables are listed in Table S6. Respondents answered these questions by selecting a value on 5-point or 7-point scales. For the radar plots, 7-point scales were transformed into 5-point scales by pooling the three central options ("Slightly disagree", "Neutral", and "Slightly agree"). Average values are reported in Table S7.

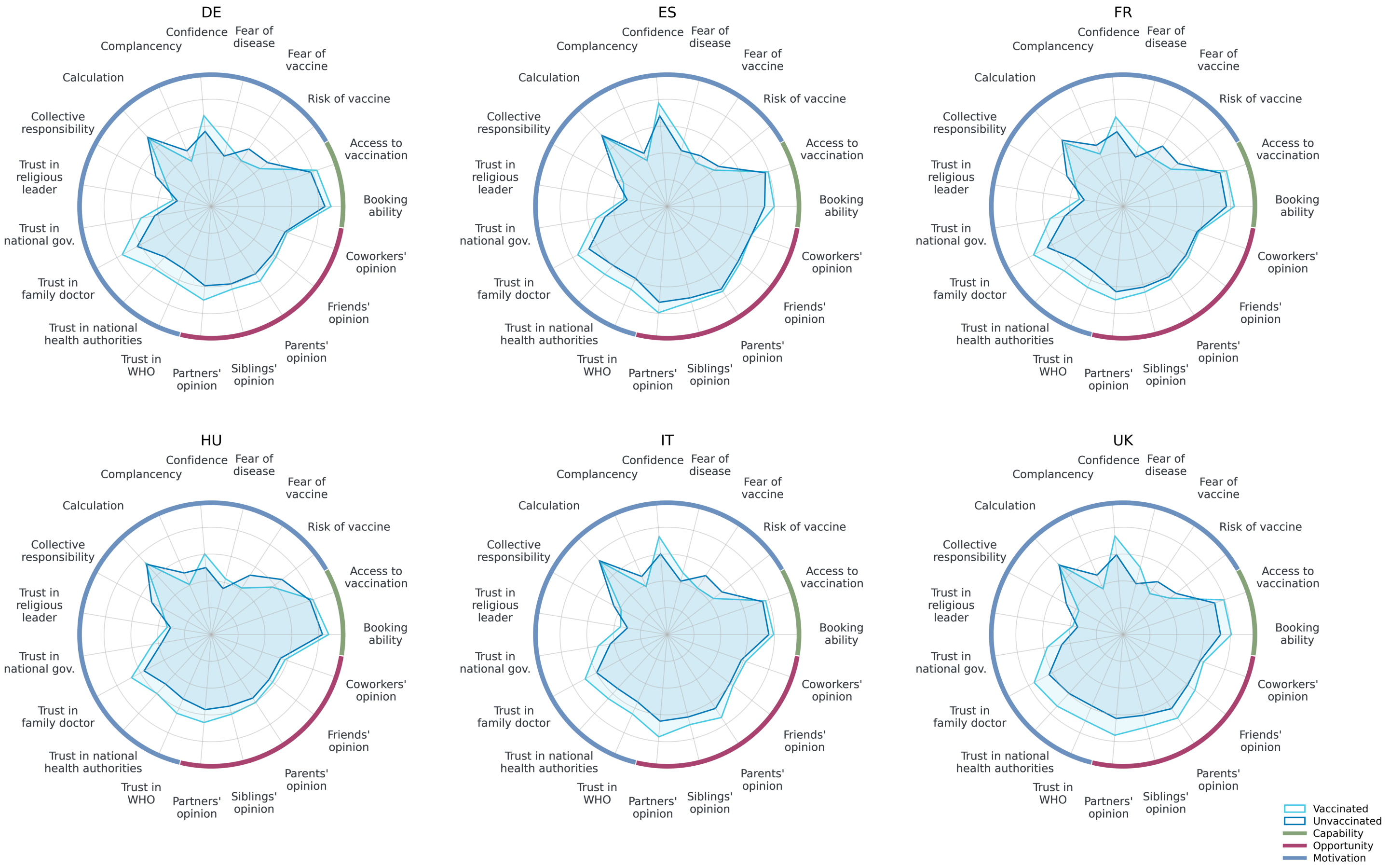

**Fig. S3** Correlation between in-person and discussion contacts Average number of daily in-person (x) and discussion (y) contacts by age group and country. The number of weekly discussion contacts as assessed in the survey was recalculated as daily. This plot excludes respondents flagged as reporting implausible contact numbers (DE n=1,452; ES n=1,725; FR n=1,395; HU n=1,561; IT n=1,780; UK n=1,180). Maximum values were capped at 30 daily contacts, with values >30 being imposed to 30.

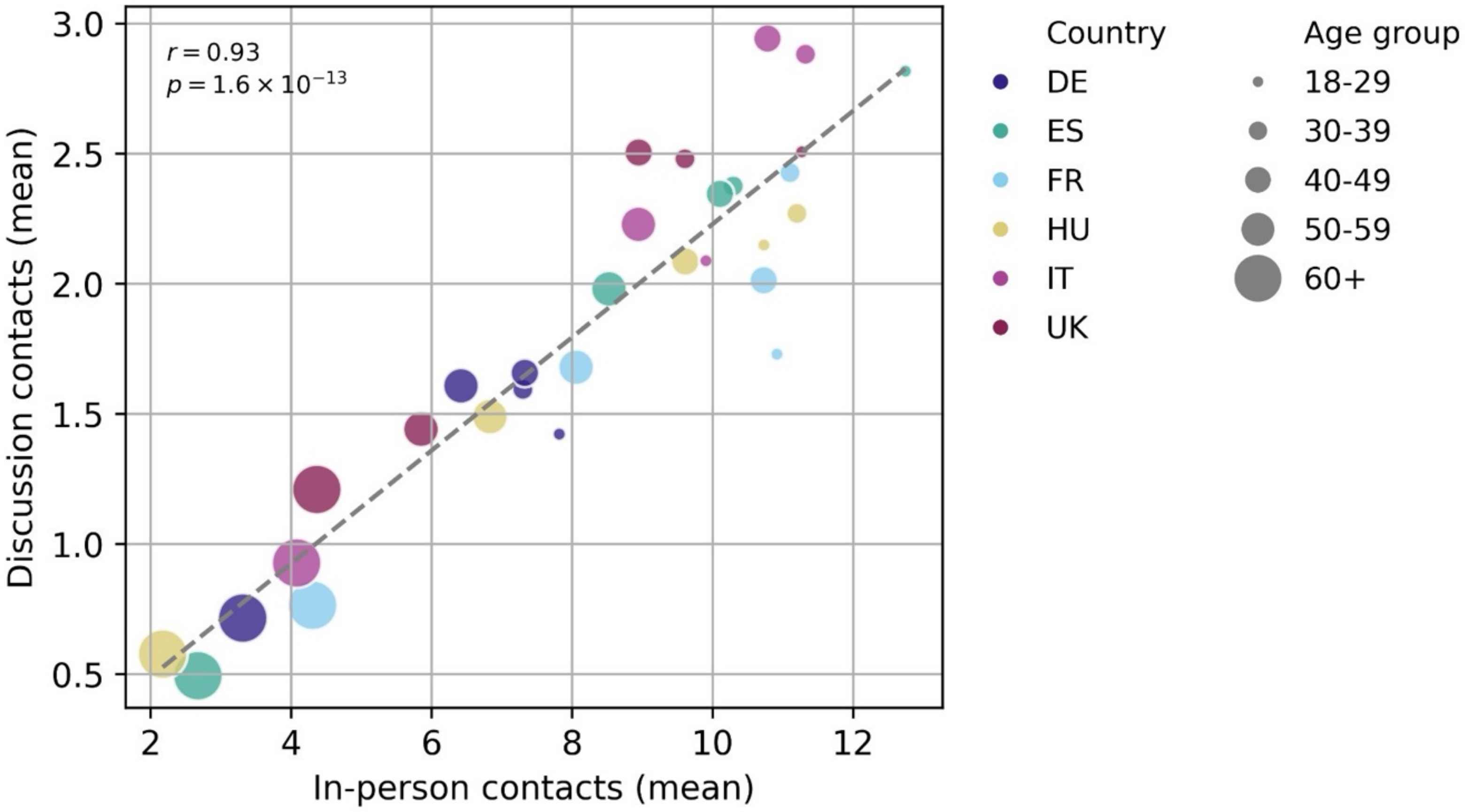

**Fig. S4** Discussion and in-person contacts by vaccination status Number of daily discussion (A) and in-person (B) contacts by age group, country, and COVID-19 vaccination status, with respondents who took up at least one dose of vaccine ever being classified as "Vaccinated". The number of weekly discussion contacts as assessed in the survey was recalculated as daily. Boxes represent medians and interquartile ranges. Whiskers represent minimum and maximum values capped at 30 daily contacts, with values >30 being imposed to 30. Small rectangles represent average values. Respondents flagged as reporting implausible contact numbers were excluded (DE n=1,452; ES n=1,725; FR n=1,395; HU n=1,561; IT n=1,780; UK n=1,180).

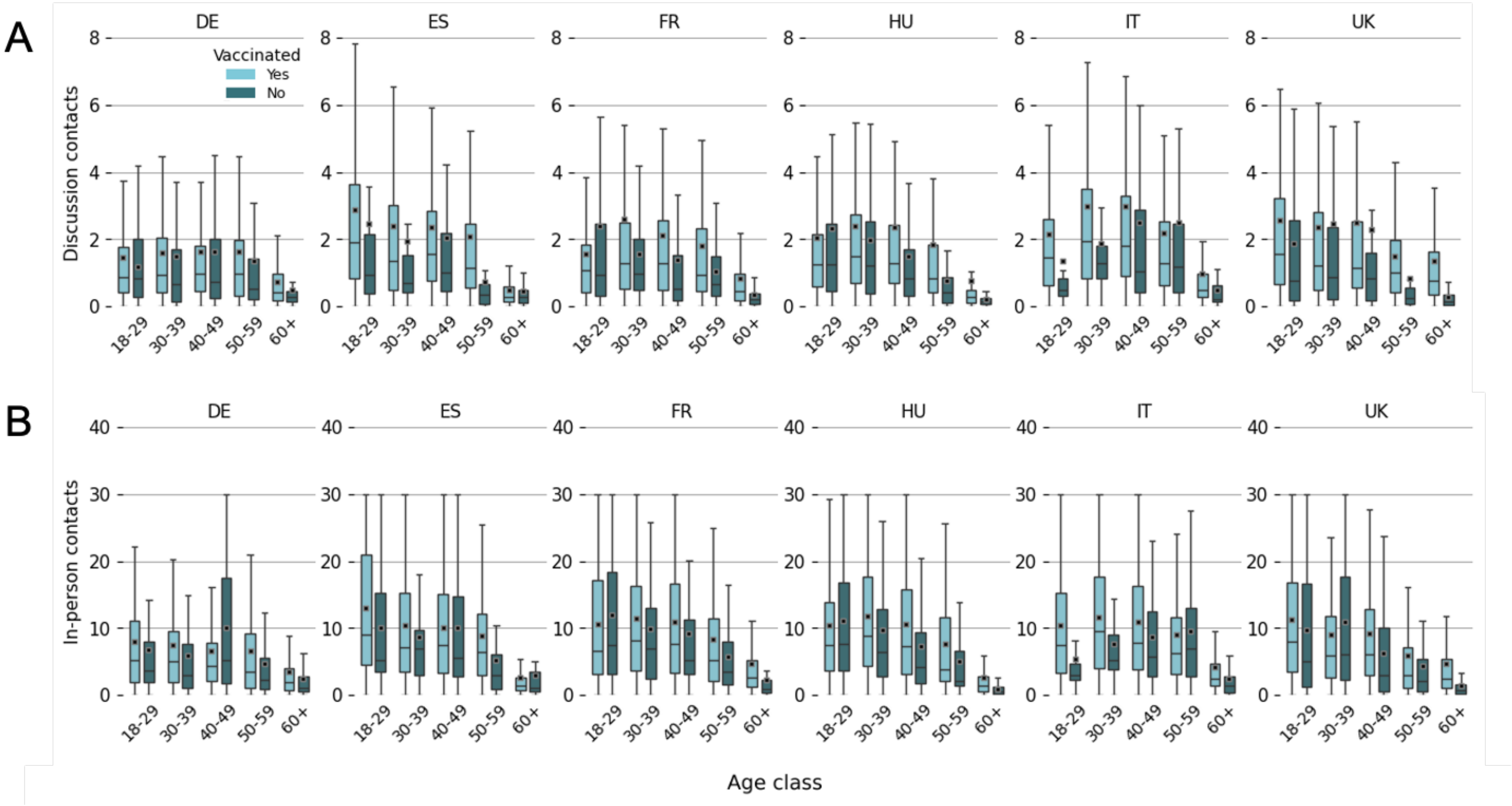

### Additional information

#### Sample size calculation

The desired number of survey participants for each country was calculated based on the required sample size to detect 5% difference in the percentage of individuals willing to vaccinate with an additional vaccine dose among non-parents versus parents, assuming this percentage in the population of parents is 50%. Setting $\alpha=0.05$ and a power of 80% ($\beta=0.2$), the required sample size is 3,317 individuals per country (1,305 parents and 2,012 non-parents).

#### Comprehension check

Before starting, please read the paragraph below and answer the following question.

MAN ARRESTED FOR STRING OF BANK THEFTS
Columbus Police have arrested a man named Bryan Simon for robbing four Ohio banks in 2018. According to court documents, Simon approached a bank counter and asked for money, claiming he had a gun. The teller gave Simon about five hundred dollars. He then told Simon he needed a driver's license to get more cash from the machine. So Simon gave the teller his license. Simon then left the bank with one thousand five hundred dollars in cash, but no license.

How much money did Simon leave the bank with?

- $500
- $1,500
- $2,000
- None of the above

#### Attention check

Among all the possible answers listed below, select the option Quite.

- Not at all
- A little
- Quite

- Very
- Completely
